# Association of Birth Weight Centiles with Infant and Child Growth Dynamics

**DOI:** 10.64898/2026.01.10.26343435

**Authors:** María Alejandra Hernandez, Richard M A Parker, Tim J Cole, Izzuddin M Aris, Henrique Barros, Johan G Eriksson, Abby F Fleisch, Barbara Heude, Yung Seng Lee, Zheyuan Li, Emily Oken, Susana Santos, Kok Hian Tan, Chloe Vainqueur, Tanja G Vrijkotte, John Wright, Tiffany C Yang, Fabian Yap, Siyu Zhou, Kate Tilling, Deborah A Lawlor, Ahmed Elhakeem

## Abstract

**Importance:** Infants born small or large for gestational age exhibit different growth patterns compared with appropriate-for-gestational age counterparts. Evidence is lacking on how birth weight centiles beyond conventional thresholds predict early life growth.

**Objective:** Quantify association of birth weight centile range with infant and child growth.

**Design:** Prospective cohort study.

**Setting:** France, Netherlands, Portugal, Singapore, United Kingdom, United States.

**Participants:** Singletons from seven birth cohort studies with repeated growth measurements from age one week to 10 years. Five European cohorts were used for discovery analysis, and remaining cohorts for replication.

**Exposures:** Birth weight centiles standardised for sex and gestational age using the INTERGROWTH-21^st^ standards and classified into deciles.

**Main Outcomes and Measures:** Infant height (*cm/month*) and weight (*g/month*) velocity at 1, 6, 12, 24 months, body mass index (BMI: *kg/m^2^*) and age (*months* or *years*) at infancy BMI peak and childhood BMI rebound, and overweight/obesity at age 10 years.

**Results:** The discovery analysis included 36,018 children (48% girls, birth years: 1991-2011). Growth velocity, peak BMI (mean: 17.7 vs. 17.3 *kg/m^2^*), age at peak BMI (10.2 vs. 10.4 *months*), rebound BMI (15.6 vs. 15.5 *kg/m^2^*), age at rebound BMI (5.3 vs. 5.0 *years*), and overweight/obesity prevalence were broadly similar in boys and girls. Compared with lowest birth weight decile (<10^th^ centile), all higher decile groups had an initially lower height velocity that reversed by 24 months, an increasingly higher weight velocity through infancy, a higher and earlier peak BMI, higher rebound BMI, and a higher probability of overweight/obesity at age 10 years. For example, mean differences in BMI and age at peak BMI, and probability of overweight/obesity at age 10 years for the 7^th^ (vs. 1^st^) birth weight decile were 0.77 *kg/m^2^* (0.72 to 0.82), -0.58 *months* (-0.69 to -0.46), and 34% (18 to 49). Birth weight was not associated with age at rebound BMI. Replication analyses (n=2,517) supported findings. Associations were typically linear and consistent for boys and girls. Deciles provided only modest predictive gains over conventional categories.

**Conclusions and Relevance:** Birth weight centiles may identify high-risk children missed by traditional thresholds, although predictive benefit over conventional groups was modest.

---

Infant and childhood growth patterns are thought to reflect developmental processes driving the risk of later obesity. For instance, rapid infant weight gain^1^, higher childhood BMI, earlier and higher BMI rebound^2^, and faster increase in childhood BMI^3^ predict higher subsequent obesity risk. Examining the predictors of these growth patterns is crucial for advancing our understanding of the developmental mechanisms underlying obesity, enabling earlier identification of at-risk children, and informing more effective interventions to reduce its burden^4,5^.

Abnormal fetal growth is an important clinical prognostic indicator of neonatal and postnatal morbidity^6^. Clinicians typically identify suboptimal fetal growth using population-based birth weight for gestational age (and sex) standards to classify infants as small for gestational age (SGA) if birth weight is below the 10^th^ centile or large for gestational age (LGA) if above the 90^th^ centile. Studies show that offspring born SGA (a possible consequence of fetal growth restriction) and LGA (often indicative of fetal overgrowth) have unique growth patterns in infancy and childhood, distinct from infants born appropriate for gestational age (AGA)^7–9^. Reliance on convenient SGA/LGA thresholds limits our understanding of how the birth weight spectrum relates to growth. Granular analyses can expand our understanding of the biology underlying obesity, and reveal subtle, and clinically relevant, growth differences, improving identification of infants at risk of later obesity^10–12^. Moreover, comparing cohorts born in different regions can improve generalisability and identify contextual factors.

The aim of this study was to quantify associations of birth weight centiles with infant height and weight growth velocity and longitudinal growth, the magnitude and timing of infant BMI peak and childhood BMI rebound, and childhood overweight/obesity in a pooled cohort, and to replicate the findings in an independent cohort.

## METHODS

### Cohort studies

Prospective pregnancy and birth cohort studies were identified from the European Union Child Cohort Network (ECCN)^13,14^ and collaborating studies. Cohorts were eligible for this study if they collected data on gestational age at birth, birth weight, and sex, and had at least five repeated measurements of length/height and weight between one week and 10 years of age. From the eligible cohorts, children were selected if they were singletons with available data on sex, gestational age, birth weight, and a minimum of two growth measurements.

Five ECCN birth cohort studies from the Netherlands (Amsterdam Born Children and their Development^15^), UK (Avon Longitudinal Study of Parents and Children^16–18^, and Born in Bradford^19^), France (Etude des Déterminants pré et post natals précoces du développement psychomoteur et de la santé de l’Enfant)^20^, and Portugal (Generation XXI)^21^ met eligibility criteria and were included (**eMethods**). Two additional birth cohorts from the USA (Project Viva)^22,23^ and Singapore (Growing Up in Singapore Towards healthy Outcomes Study)^24^ were also included (**eMethods**). Mothers were recruited during pregnancy (six studies) or labour (Generation XXI). All studies collected information from parents and their offspring using questionnaires, medical or health records, and dedicated research clinics. All studies had ethical approval from their local or national ethics committees and study participants gave informed consent/assent to participate in respective cohorts and secondary analyses (**eMethods**). The five European birth cohorts were combined and used as the Discovery cohort. The two additional cohorts from the USA and Singapore were combined to create the Replication cohort, which was used to validate the findings in the Discovery cohort.

### Birthweight centiles

Gestational age at birth and birth weight were retrieved from health records. Birth weight was standardised by sex and gestational age using the International Fetal and Newborn Growth Consortium for 21^st^ Century (INTERGROWTH-21^st^) standards. The INTERGROWTH-21^st^ standards were developed on >20,000 healthy live births from eight countries (Brazil, Italy, Oman, United Kingdom, United States, China, India, Kenya)^25^. For our main analysis, we categorised standardised birth weight into 10 centile groups which we refer to as decile groups throughout the paper. We used the first decile (<10^th^ centile) as referent group.

### Infant and child growth outcomes

Repeated length/height and weight measurements from age one week to 10 years, and the ages of measurement, were available from health records and research clinic assessments in each cohort (**eFigure 1**). Data were cleaned to remove errors^26^. BMI was calculated as weight (kg) divided by height (m) squared. Median (interquartile range) number of measurements per child was 7 (8) in the Discovery Cohort and 14 (8) in the Replication Cohort. Height, weight, and BMI trajectories up to age 10 years were estimated using sex-stratified P-splines linear mixed effects models^27,28^ separately in the Discovery and Replication cohorts (**eMethods**).

The fitted growth curves (**eFigure 2**) were used to estimate growth outcomes for subsequent analysis. Growth rate during infancy was estimated by using derivatives of the fitted height and weight trajectories to obtain growth velocity at ages 1, 6, 12, and 24 months. To examine growth patterns, we used the fitted trajectories to predict height and weight values at 3-month intervals from age 1-60 months and converted these to height-for-age and weight-for-age Z-scores using the WHO Child Growth Standards. Magnitude (BMI) and timing (age) of infant BMI peak and childhood BMI rebound were calculated from the derivatives of the fitted BMI trajectories. The prevalence of overweight or obesity at age 10 years was derived by applying the International Obesity Task Force age- and sex-specific cut-off points to predicted BMIs^29^. Further details are in the **eMethods**.

### Statistical analysis

Mean differences in infant growth velocity (at ages 1, 6, 12, and 24 months) and BMI and age at infant peak BMI and childhood rebound BMI for each birth weight decile group (vs. first decile) were estimated using linear regression. Height-for-age and weight-for-age Z-score trajectories for each birth weight decile group from 1 month to 5 years were estimated using linear mixed effects models. Logistic regression was used to estimate mean differences in probability of overweight or obesity at age 10 years for each birth weight decile group (vs. first decile). All models were fitted in males and females combined, and separately in the Discovery and Replication cohorts with adjustment made for sex and birth cohort. Z-score trajectory models included a natural spline for age^30^ plus its interaction with birth weight decile group. Meta-regression was used to test for differences in estimates between the Discovery and Replication cohort.

The following additional analyses were conducted in the Discovery cohort. Sex differences were examined by fitting models with interactions between birth weight decile group and sex. To evaluate whether using birth weight decile groups offered predictive benefit over the three conventional groups (SGA, AGA, LGA), we used the coefficient of determination (R^2^) and the area under the receiver operating characteristics curve to compare the performance of two competing linear and logistic models, respectively. We interrogated the nonlinear nature of the associations by comparing models with birth weight Z-score entered as a linear term and as natural spline terms. Lastly, because prenatal growth could influence postnatal growth^31,32^, we explored robustness of our results to measured confounders (factors that could influence both prenatal and postnatal growth) by fitting models with further adjustment for pregnancy-related covariates (maternal parity, maternal ethnicity, and both parents’ BMI, age, smoking, and education). Missing covariate data were imputed using multiple imputation, and results were compared with complete case analysis. All analysis was done using R 4.5.2 (R Project for Statistical Computing). Further details on analyses are in the **eMethods**.

## RESULTS

### Participant Characteristics

A total of 36,018 births (48% girls) and 2,517 births (51% girls) were included in Discovery and Replication cohorts, respectively (**Figure 1**). Discovery cohort participants were born in 1991-2011 in Amsterdam, the Netherlands (16%), Bristol, UK (34%), Bradford, UK (29%), Nancy and Poitiers, France (5%), and Porto, Portugal (16%). Replication cohort participants were born in 1999-2010 in Massachusetts, USA (64%) and Singapore (36%).

**Figure 1.**
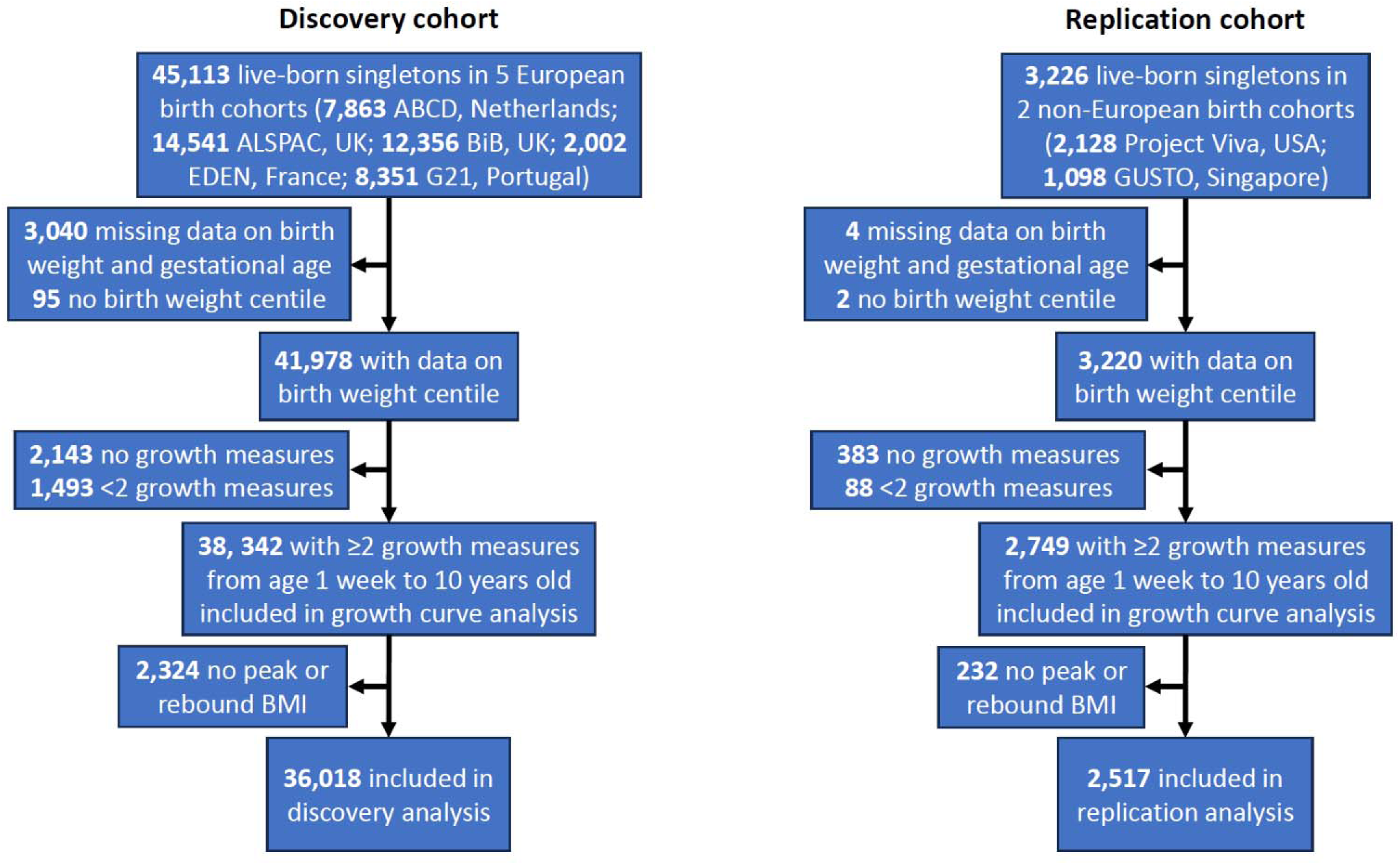
Study flowchart.

Birth characteristics and growth outcomes are summarised in **Table 1** for the Discovery and Replication cohorts, and in **eTable 1** for each sub-cohort. Mean birth weight, gestational age, and birth weight centile were similar in both cohorts, including a similar range of gestational age (25 to 43 *weeks*). In both cohorts, the lowest four decile groups included less than 10% of study participants whereas the top two groups had more than 10%. Infant growth velocity and peak BMI, and the BMI and age at rebound BMI were broadly similar in both cohorts but the Discovery cohort had an older age at peak BMI and a lower prevalence of overweight/obesity at age 10 years. Inter correlations of the continuous growth outcomes were similar for both cohorts and included moderate positive associations between peak and rebound BMI, and negligible associations between the ages of peak and rebound BMI (**eFigure 3**).

**Table 1.**
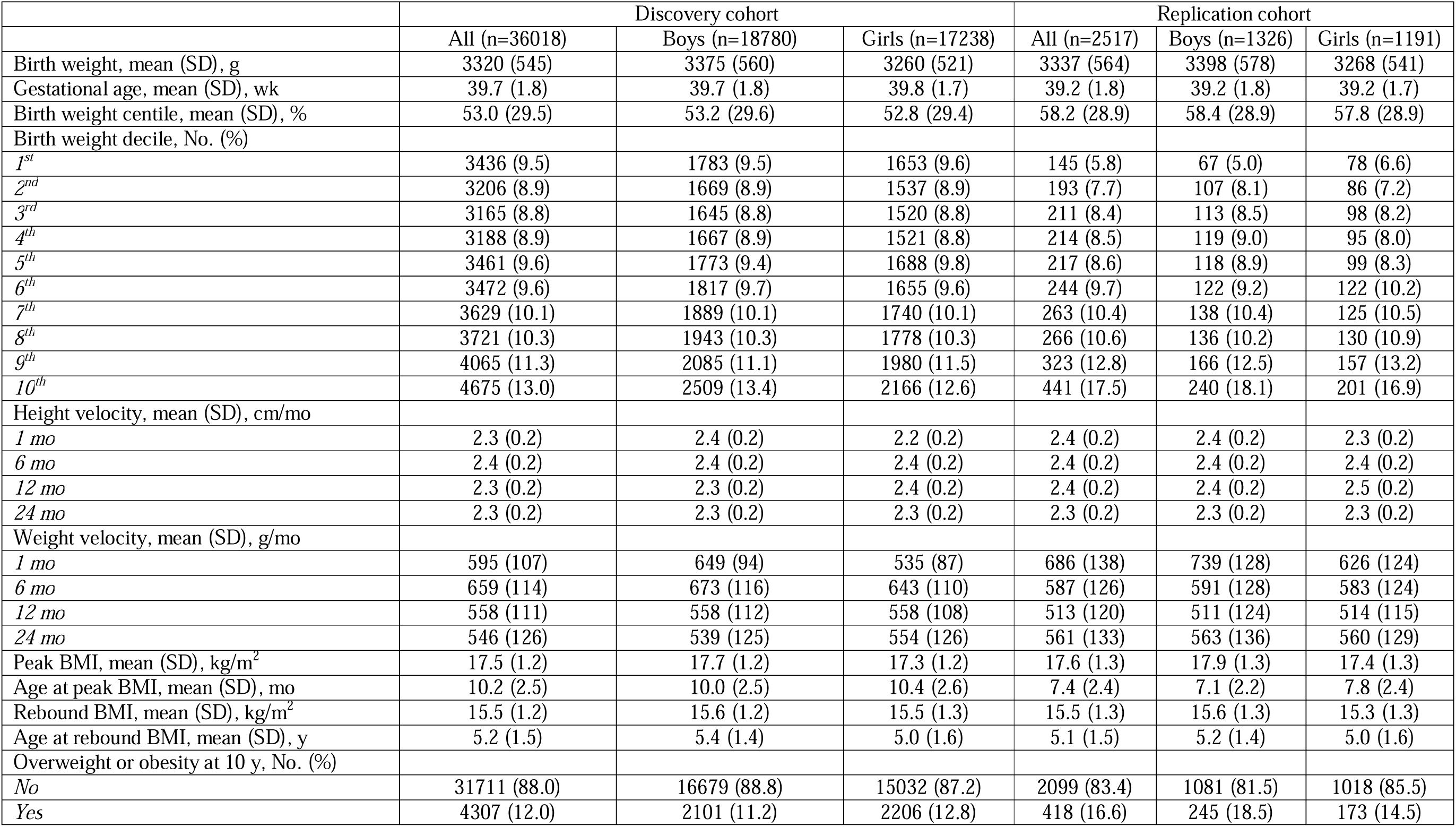
Birth characteristics and infant/child growth outcomes in the discovery and replication cohorts. Discovery cohort was born 1991-2011; Replication cohort was born 1999-2010. Birth weight was standardised for sex and gestational using INTERGROWTH-21^st^ standards. Decile 1: <10^th^, decile 2: 10^th^ to <20^th^, decile 3: 20^th^ to <30^th^, decile 4: 30^th^ to <40^th^, decile 5: 40^th^ to <50^th^, decile 6: 50^th^ to <60^th^, decile 7: 60^th^ to <70^th^, decile 8: 70^th^ to <80^th^, decile 9: 80^th^ to 90^th^, decile 10: >90^th^.

|  | Discovery cohort |  |  | Replication cohort |  |  |
| --- | --- | --- | --- | --- | --- | --- |
|  | All (n=36018) | Boys (n=18780) | Girls (n=17238) | All (n=2517) | Boys (n=1326) | Girls (n=1191) |
| Birth weight, mean (SD), g | 3320 (545) | 3375 (560) | 3260 (521) | 3337 (564) | 3398 (578) | 3268 (541) |
| Gestational age, mean (SD), wk | 39.7 (1.8) | 39.7 (1.8) | 39.8 (1.7) | 39.2 (1.8) | 39.2 (1.8) | 39.2 (1.7) |
| Birth weight centile, mean (SD), % | 53.0 (29.5) | 53.2 (29.6) | 52.8 (29.4) | 58.2 (28.9) | 58.4 (28.9) | 57.8 (28.9) |
| Birth weight decile, No. (%) |  |  |  |  |  |  |
| 1<sup>st</sup> | 3436 (9.5) | 1783 (9.5) | 1653 (9.6) | 145 (5.8) | 67 (5.0) | 78 (6.6) |
| 2<sup>nd</sup> | 3206 (8.9) | 1669 (8.9) | 1537 (8.9) | 193 (7.7) | 107 (8.1) | 86 (7.2) |
| 3<sup>rd</sup> | 3165 (8.8) | 1645 (8.8) | 1520 (8.8) | 211 (8.4) | 113 (8.5) | 98 (8.2) |
| 4<sup>th</sup> | 3188 (8.9) | 1667 (8.9) | 1521 (8.8) | 214 (8.5) | 119 (9.0) | 95 (8.0) |
| 5<sup>th</sup> | 3461 (9.6) | 1773 (9.4) | 1688 (9.8) | 217 (8.6) | 118 (8.9) | 99 (8.3) |
| 6<sup>th</sup> | 3472 (9.6) | 1817 (9.7) | 1655 (9.6) | 244 (9.7) | 122 (9.2) | 122 (10.2) |
| 7<sup>th</sup> | 3629 (10.1) | 1889 (10.1) | 1740 (10.1) | 263 (10.4) | 138 (10.4) | 125 (10.5) |
| 8<sup>th</sup> | 3721 (10.3) | 1943 (10.3) | 1778 (10.3) | 266 (10.6) | 136 (10.2) | 130 (10.9) |
| 9<sup>th</sup> | 4065 (11.3) | 2085 (11.1) | 1980 (11.5) | 323 (12.8) | 166 (12.5) | 157 (13.2) |
| 10<sup>th</sup> | 4675 (13.0) | 2509 (13.4) | 2166 (12.6) | 441 (17.5) | 240 (18.1) | 201 (16.9) |
| Height velocity, mean (SD), cm/mo |  |  |  |  |  |  |
| 1 mo | 2.3 (0.2) | 2.4 (0.2) | 2.2 (0.2) | 2.4 (0.2) | 2.4 (0.2) | 2.3 (0.2) |
| 6 mo | 2.4 (0.2) | 2.4 (0.2) | 2.4 (0.2) | 2.4 (0.2) | 2.4 (0.2) | 2.4 (0.2) |
| 12 mo | 2.3 (0.2) | 2.3 (0.2) | 2.4 (0.2) | 2.4 (0.2) | 2.4 (0.2) | 2.5 (0.2) |
| 24 mo | 2.3 (0.2) | 2.3 (0.2) | 2.3 (0.2) | 2.3 (0.2) | 2.3 (0.2) | 2.3 (0.2) |
| Weight velocity, mean (SD), g/mo |  |  |  |  |  |  |
| 1 mo | 595 (107) | 649 (94) | 535 (87) | 686 (138) | 739 (128) | 626 (124) |
| 6 mo | 659 (114) | 673 (116) | 643 (110) | 587 (126) | 591 (128) | 583 (124) |
| 12 mo | 558 (111) | 558 (112) | 558 (108) | 513 (120) | 511 (124) | 514 (115) |
| 24 mo | 546 (126) | 539 (125) | 554 (126) | 561 (133) | 563 (136) | 560 (129) |
| Peak BMI, mean (SD), kg/m<sup>2</sup> | 17.5 (1.2) | 17.7 (1.2) | 17.3 (1.2) | 17.6 (1.3) | 17.9 (1.3) | 17.4 (1.3) |
| Age at peak BMI, mean (SD), mo | 10.2 (2.5) | 10.0 (2.5) | 10.4 (2.6) | 7.4 (2.4) | 7.1 (2.2) | 7.8 (2.4) |
| Rebound BMI, mean (SD), kg/m<sup>2</sup> | 15.5 (1.2) | 15.6 (1.2) | 15.5 (1.3) | 15.5 (1.3) | 15.6 (1.3) | 15.3 (1.3) |
| Age at rebound BMI, mean (SD), y | 5.2 (1.5) | 5.4 (1.4) | 5.0 (1.6) | 5.1 (1.5) | 5.2 (1.4) | 5.0 (1.6) |
| Overweight or obesity at 10 y, No. (%) |  |  |  |  |  |  |
| No | 31711 (88.0) | 16679 (88.8) | 15032 (87.2) | 2099 (83.4) | 1081 (81.5) | 1018 (85.5) |
| Yes | 4307 (12.0) | 2101 (11.2) | 2206 (12.8) | 418 (16.6) | 245 (18.5) | 173 (14.5) |

### Association with infant height and weight growth velocity

In the Discovery cohort, higher birth weight decile was associated with a lower infant height velocity at age 1 month (**Figure 2**). This association became weaker with increasing age and by 24 months had reversed to an association with higher height velocity. Higher birth weight decile was associated with higher infant weight velocity, and this association became stronger with increasing age (**Figure 2**). For example, mean differences in height and weight velocity at age 6 months for the 7^th^ (vs. 1^st^) birth weight decile were -0.06 *cm/mo* (95%CI: -0.07 to - 0.05) and 21.9 *g/mo* (16.7 to 27.2). Effect sizes increased with higher decile groups. Results for height and weight velocity at 6 months and weight velocity at other ages were generally similar in the Replication cohort. The Replication cohort showed stronger associations with height velocity at age 1 month (*P* _cohort difference_ = 0.03) and 24 months (*P* _cohort difference_ = 0.002) and mainly null association with height velocity at 12 months (*P* _cohort difference_ = 0.00002).

**Figure 2.**
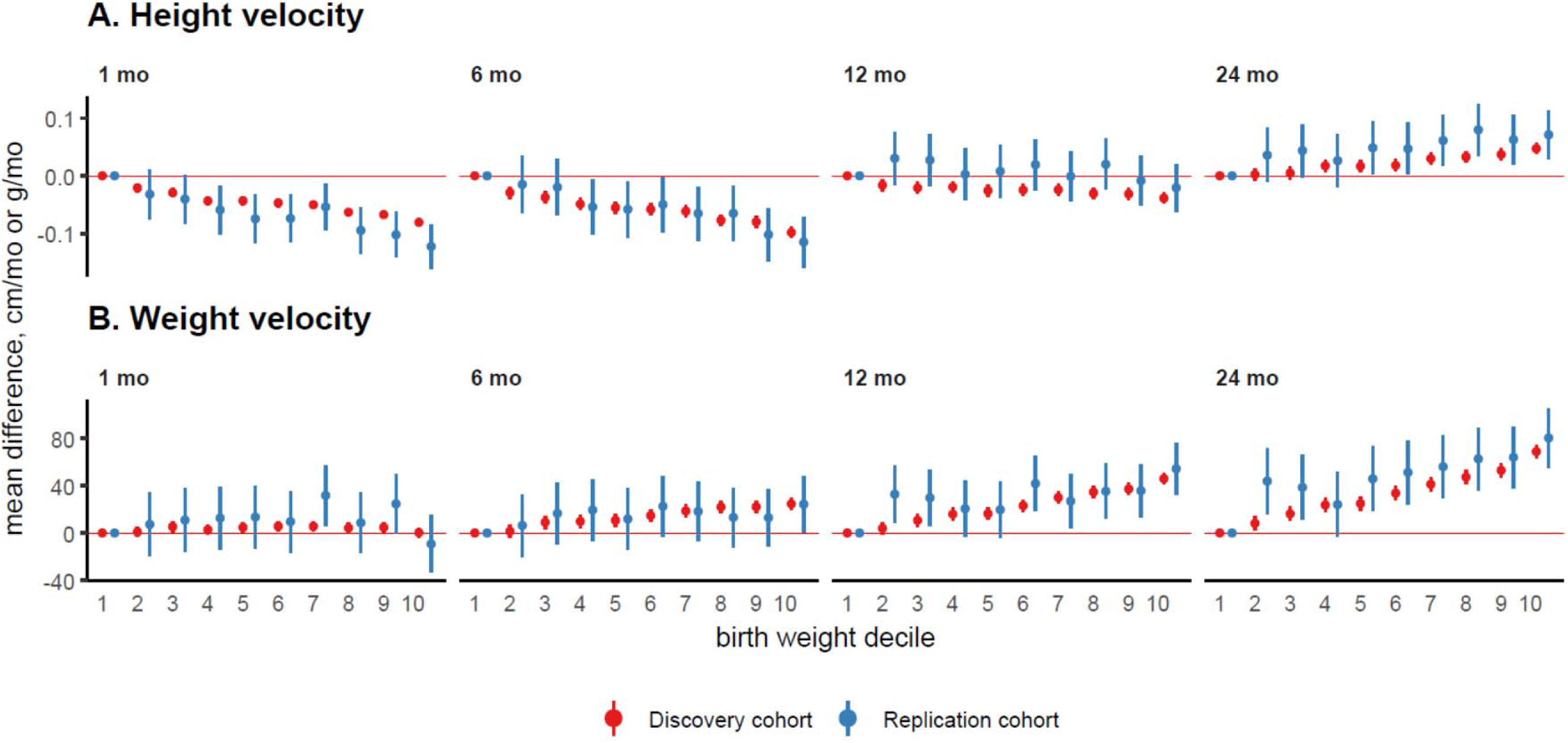
Mean difference in infant height and weight growth velocity at age 1, 6, 12, and 24 months. Figure shows mean differences (points) and 95% confidence intervals (vertical bars) in infant height and weight growth velocity at ages 1, 6, 12 and 24 months for each birth weight decile group (vs. <1^st^ decile). Models were fitted separately in the discovery and replication cohorts and adjusted for sex and cohort. Decile 1: <10^th^, decile 2: 10^th^ to <20^th^, decile 3: 20^th^ to <30^th^, decile 4: 30^th^ to <40^th^, decile 5: 40^th^ to <50^th^, decile 6: 50^th^ to <60^th^, decile 7: 60^th^ to <70^th^, decile 8: 70^th^ to <80^th^, decile 9: 80^th^ to 90^th^, decile 10: >90^th^. Numerical results are in eTable 2.

### Association with height and weight growth from 1 month to 5 years

In the Discovery cohort, mean height and weight Z scores were higher in higher birth weight decile groups. The magnitude of the differences between birth weight groups was largest at the earliest age and progressively attenuated with age (**Figure 3**). For height, children in the lowest three and highest three birth weight decile groups respectively remained below and above the WHO average height up to age 5 years. For weight, only the lowest decile group and highest two decile groups respectively remained below and above the WHO average. The Replication cohort showed consistent results for both height (*P* _cohort difference_ = 0.9) and weight (*P* _cohort difference_ = 0.4).

**Figure 3.**
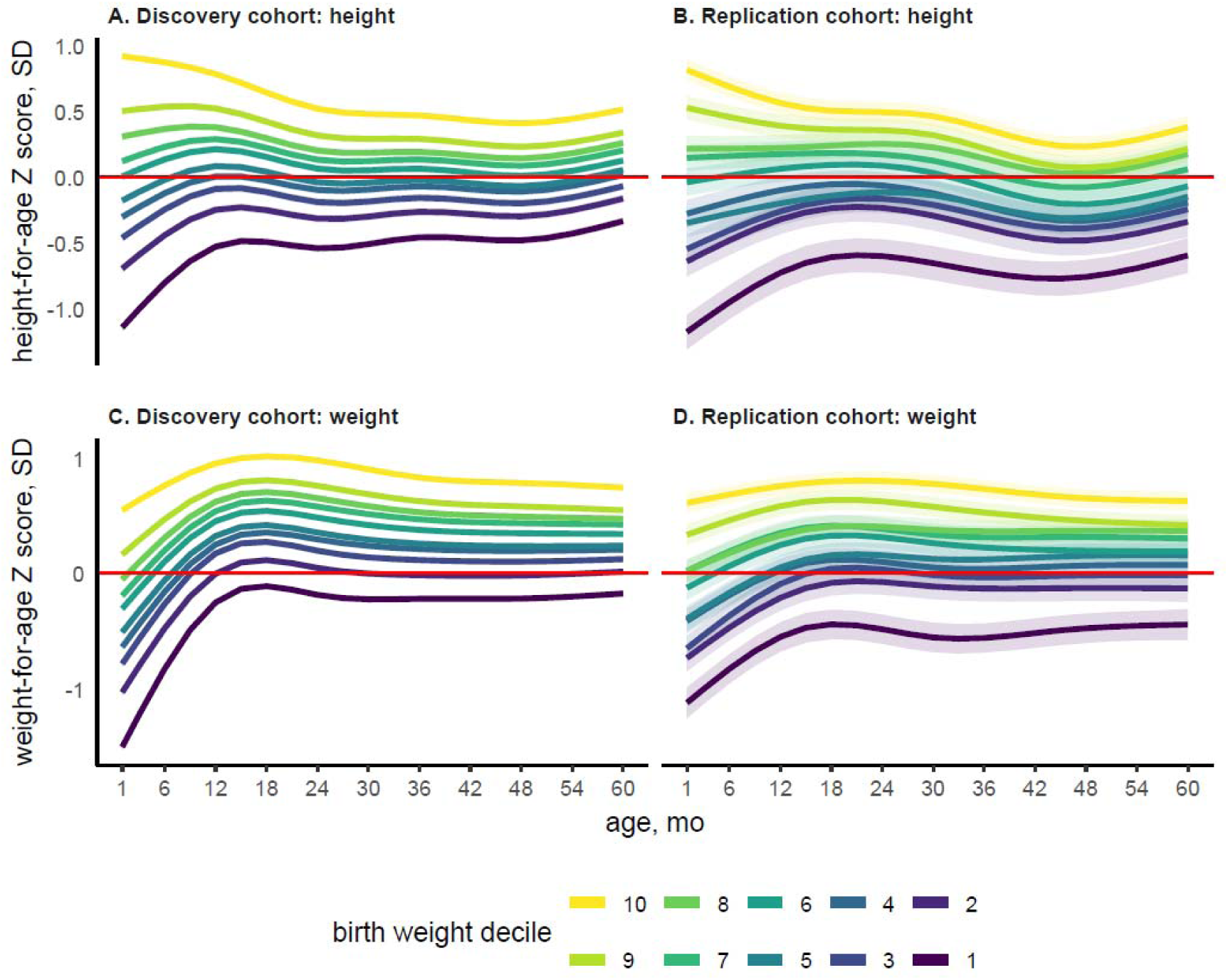
Mean height and weight Z score trajectory from age 1 month to 5 years. Figure shows mean (lines) and 95% confidence intervals (bands) for WHO height-for-age and weight-for-age Z-score trajectory from 1 month to 5 years for each birth weight decile group. Models were fitted separately in the Discovery and Replication cohort. Models were adjusted for sex and cohort and included a natural spline for age plus its interaction with birth weight. Decile 1: <10^th^, decile 2: 10^th^ to <20^th^, decile 3: 20^th^ to <30^th^, decile 4: 30^th^ to <40^th^, decile 5: 40^th^ to <50^th^, decile 6: 50^th^ to <60^th^, decile 7: 60^th^ to <70^th^, decile 8: 70^th^ to <80^th^, decile 9: 80^th^ to 90^th^, decile 10: >90^th^

### Association with infant peak BMI and childhood rebound BMI

In the Discovery cohort, higher birth weight groups were associated with higher peak and rebound BMI, and younger age at peak, but not rebound, BMI (**Figure 4**). For example, the mean differences in BMI and age at infant peak BMI for the 7^th^ (vs. 1^st^) birth weight decile were 0.77 *kg/m^2^* (0.72 to 0.82) and -0.58 *months* (-0.69 to -0.46). Effect sizes increased with higher decile groups. The Replication cohort showed consistent results for peak and rebound BMI (*P* _cohort difference_ = 0.7 and 0.8 respectively), a weaker association with age at peak BMI (*P* _cohort difference_ = 0.001) and possible U-shaped association with age at rebound BMI (*P* _cohort difference_ = 0.008).

**Figure 4.**
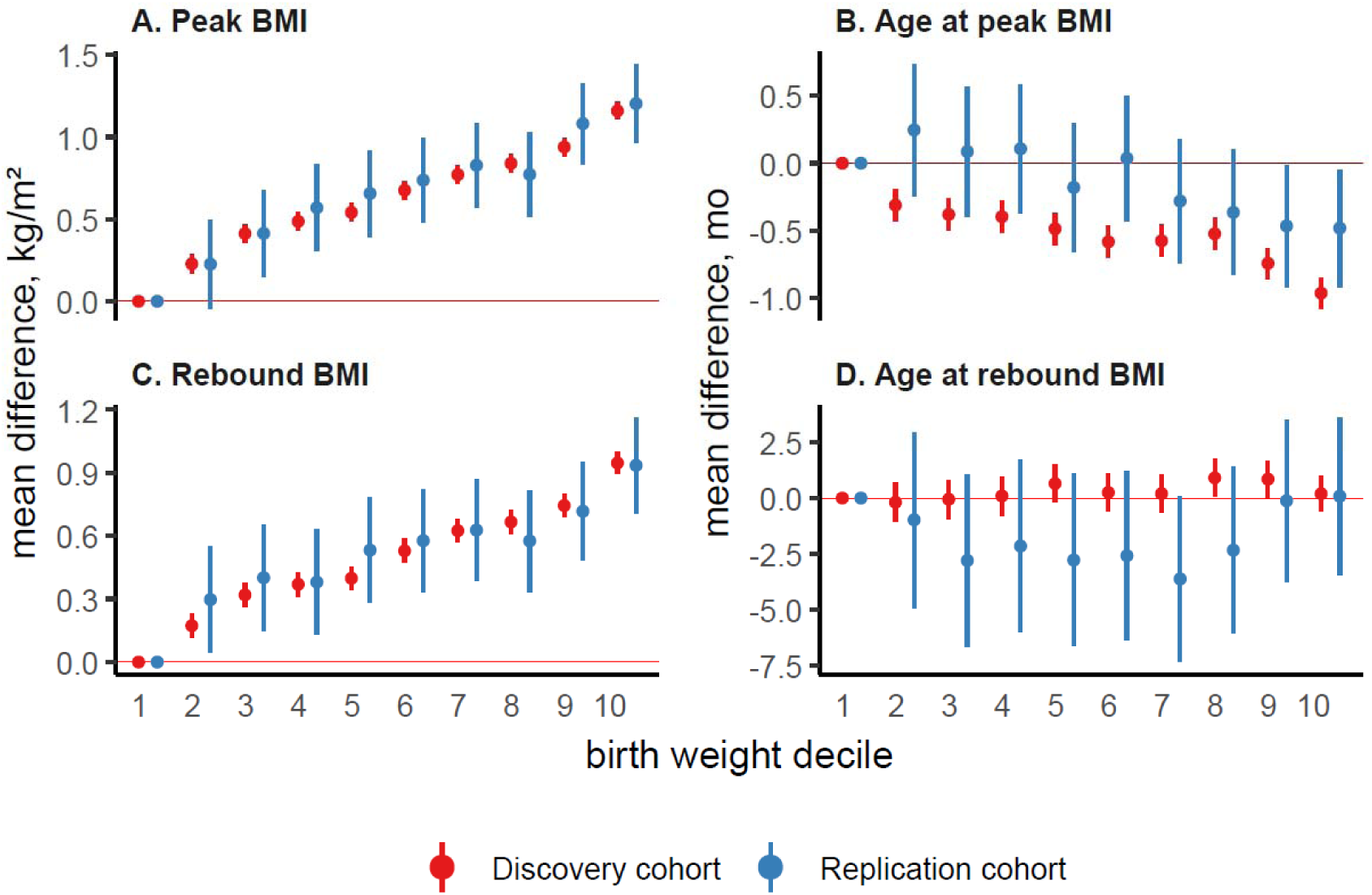
Mean difference in magnitude and timing of infant peak BMI and childhood rebound BMI. Figure shows mean differences (points) and 95% confidence intervals (vertical bars) in BMI and age at infant peak BMI and childhood rebound BMI for each birth weight decile category (vs. <10^th^ birth weight centile). Models were fitted separately in the European and replication (non-European) birth cohort and adjusted for sex and cohort. Decile 1: <10^th^, decile 2: 10^th^ to <20^th^, decile 3: 20^th^ to <30^th^, decile 4: 30^th^ to <40^th^, decile 5: 40^th^ to <50^th^, decile 6: 50^th^ to <60^th^, decile 7: 60^th^ to <70^th^, decile 8: 70^th^ to <80^th^, decile 9: 80^th^ to 90^th^, decile 10: >90^th^. Numerical results are presented in eTable 2.

### Association with overweight or obesity at age 10 years

In the Discovery cohort, higher birth weight groups were associated with increasingly higher probability of overweight or obesity at 10 years (**eTable 2**). For example, mean difference in probability of overweight/obesity for the 7^th^ (vs. 1^st^) birth weight decile was 34% (18 to 49). This analysis was not done in the Replication cohort due to low numbers; however, treating birth weight decile groups as continuous showed that a per category increase in birth weight decile associated with 9% (5 to 13) higher probability of overweight/obesity. The equivalent estimate in the Discovery cohort (7%, 6 to 9) was consistent with the Replication cohort’s estimate (*P* cohort difference = 0.6).

### Additional (Discovery cohort) results

For most outcomes, there was no evidence of a sex difference in associations. Differences in height and weight Z-score trajectories by birth weight decile appeared larger in girls vs. boys (*P* _sex difference_ = 0.0005 and 0.003 respectively) (**eFigure 4**). Using birth weight decile groups provided only modest improvements in predictive performance vs. using three conventional groups; the largest improvement was for weight Z-score trajectories (Δ*R^2^* +4.3%). Comparing linear vs. spline terms for birth weight Z-score identified linear associations with most growth outcomes, subtle nonlinear associations with height velocity at age 1 month and 6 months and overweight/obesity, and an inverse U-shaped association with weight velocity at age 1 month (**eFigure 5**). Lastly, association estimates were only slightly attenuated after adjustment for confounders (**eTables 3-4**).

## DISCUSSION

We used data from >36,000 European and >2,500 non-European singletons born 1991-2011 to quantify and replicate the associations between birth weight centiles and infant and child growth features. Higher birth weight decile was initially associated with lower infant height velocity, but the association reversed at age 24 months. Higher birth weight was associated with higher infant weight velocity with this association strengthening through infancy. Notwithstanding differences in infancy growth velocity, infants in the lowest birth weight decile remained below WHO average height and weight up to age 5 years, while infants in the highest group consistently exceeded them. Higher birth weight associated with higher peak and rebound BMI, an earlier peak (but not rebound) BMI, and higher probability of overweight/obesity at age 10 years. Results were replicated in an independent cohort and were consistent in males and females and mainly linear in nature.

Our findings corroborate and expand on previous studies that have examined association of conventional birth weight for gestational age groups or continuous birth weight scores with early life growth outcomes. The replication results from our pooled US and Singapore birth cohorts demonstrate consistency in direction and substantial consistency in magnitude with associations in our pooled European (Discovery) birth cohort, and align with prior studies in Europe, Australia, New Zealand, and China^7–9,33–41^.

Our finding of a continuum of increasing differences in growth outcomes with higher birth weight groups was corroborated by spline analysis showing mostly linear associations, which included, interestingly, a similar linear trend for peak and rebound BMI. Taken together, our results support the use of birth weight centiles as a predictive tool for infant and childhood growth across diverse settings, including in boys and girls, given our largely consistent results by sex. Nonetheless, there was only a modest predictive benefit of using birth weight deciles over conventional groups (i.e., LGA, SGA, AGA), indicating that the latter also continue to be useful for detecting early postnatal growth differences. However, our findings indicate that clinicians should be aware that infants below the LGA threshold may still be at elevated risk for adverse growth patterns and subsequent overweight or obesity.

Infant and childhood growth patterns can be conceptualised as a continuation of intrauterine growth trajectories, which are demonstrably linked to birth anthropometrics. Genetic factors are likely to explain some of our findings, particularly for growth outcomes beyond infancy where perinatal factors likely play a lesser role^42–45^. Future studies could explore the distinct contributions of maternal and paternal genetic factors in fetal and postnatal growth. Growth patterns may also be caused in part by intrauterine environments^31,32^. For instance, the accelerated linear growth in early infancy in the lowest birth weight group could be a compensatory response to intrauterine growth restriction^46,47^. Our observation that the associations remained robust to adjustment for measured confounders supports this hypothesis, however this exploratory analysis requires a more thorough study and triangulation.

### Strengths and limitations

Strengths of this study include its large sample size, which enabled granular analyses of the birth weight centile range including exploration of nonlinear effects and sex differences. The use of an independent pooled cohort study for replication supports the validity of our results and their generalisability. Our analyses of growth outcomes that reflect early developmental processes to obesity risk provides results with relevance to public health. Our use of P-splines allowed for complex nonlinear growth to be modelled flexibly, with penalization guarding against overfitting. We excluded children with missing birth weight or gestational age data, children with fewer than two repeated growth measurements, and those with no identifiable BMI peak or rebound, which may have reduced precision of our estimates. The participants were born 15-35 years ago in high-income countries in Europe, the USA, and Singapore, and so the findings may not generalise to more recent birth cohorts or to those in lower-income settings. In keeping with previous studies, we used the conventional formulation for BMI appropriate for adults, but this may not always be optimal for children^48^. Lastly, we did not directly measure fetal growth or postnatal fat mass and therefore we are unable to determine the growth trajectories preceding birth weight or differences in postnatal body composition.

### Conclusions

Higher birth weight for gestational age across the centile distribution was associated with key infant and child growth features underlying early developmental processes for risks of higher adiposity. Although decile-based birth weight classifications provided only modest predictive gains beyond conventional size for gestational age thresholds, our findings suggest that the range used to define appropriate birth weight may be too wide. The robustness of predictions across populations and sex, together with their linear associations, indicates that birth weight centile position could be routinely integrated into clinical practice as a supplementary, more granular, screening tool to identify infants at higher risk for suboptimal growth patterns and obesity who are missed by conventional cut-offs. These findings also further underscore the importance of early intervention, including antenatally, to improve early life growth. Future research could examine the predictive value of birth weight centile for subsequent pubertal growth characteristics and develop more sophisticated postnatal growth prediction models that combine birth weight centile with other established early life factors.

## Data Availability

Data used in this study can be accessed by making an application to each individual cohort study.

## ACKNOWLEDGEMENTS

The research leading to these results has received funding from the European Research Council under grant agreement No 101021566 (ART-HEALTH), the EU Horizon 2020 research and innovation program under grant agreements 733206 (LifeCycle) and 874739 (LongITools), the UK Medical Research Council (MC_UU_00032/2, MC_UU_00032/5, MC_UU_00011/6, UKRI481), and University of Bristol Elizabeth Blackwell Institute for Health Research Institutional Strategic Support Fund. None of the funders influenced the study design, reporting or interpretation of results. The views expressed in this paper are those of the authors and not necessarily any acknowledged funder. All authors declare no competing interests. AE had full access to the data in the study and takes responsibility for the integrity of the data and the accuracy of the data analysis. Cohort-specific acknowledgements and funding statements are provided in the supplement (eText 1).

