## Supplementary Material for "Association of Birth Weight Centiles with Infant and Child Growth Dynamics"

*María Alejandra Hernandez PhD^1,2^, Richard M A Parker PhD^1,2^,* *Tim J Cole ScD^3^, Izzuddin M Aris PhD^4^, Henrique Barros PhD^5^, Johan G Eriksson MD^6,7^, Abby F Fleisch MD, MPH^8,9^, Barbara Heude PhD^10^, Yung Seng Lee PhD^6,7^, Zheyuan Li PhD^11^, Emily Oken MD^4^, Susana Santos PhD^5^, Kok Hian Tan MBBS, MMed^12^, Chloe Vainqueur PhD^10^, Tanja G Vrijkotte PhD^13^, John Wright MD^14^, Tiffany C Yang PhD^14^, Fabian Yap MD^12^, Siyu Zhou MD^13^, Kate Tilling PhD^1,2^, Deborah A Lawlor PhD^1,2^, Ahmed Elhakeem PhD^1,2^*

### eMethods

1. ***Discovery cohort***
   1.1 ***ABCD cohort description***

All pregnant women living in Amsterdam, Netherlands, between 2003 and 2004 were invited to participate in the ABCD study during their first prenatal visit to an obstetric care provider (general practitioner, midwife, or gynaecologist)^1^. Of the 12,373 women approached, 8,266 women were recruited and filled out a pregnancy questionnaire. Of the 8,266 respondents, 7,863 women gave birth to viable singleton infants, and 132 women gave birth to viable multiples. From this group, 7,050 granted permissions for follow-up, and 7,043 granted permissions for perusal of her and her child’s medical files.

Gestational age (in days) and birth weight (in grams) were retrieved from Youth Health Care registration of the Public Health Service in ABCD^2^. Length/height and weight were obtained from the Youth Health Care registration of the Public Health Service in Amsterdam. Trained nurses measured length/height and weight (following a standard procedure) at regular follow-ups between birth and age 10 years. The entire cohort were also invited to a research clinic at age 5 years where heights and weights were measured by trained staff. Of the 7,863 singleton live births, 6,384 had data on gestational age at birth and birth weight, and of these, 6,366 had data on birth weight centile. After cleaning growth data and excluding offspring with fewer than two growth measurements between age 1 week and 10 years, there was a total of 5,886 offspring left for growth curve modelling.

Approval for ABCD was obtained from the Central Committee on Research involving Human Subjects in the Netherlands, the Medical Ethical Committees of the participating hospitals, and from the Registration Committee of the Municipality of Amsterdam. Written informed consent was obtained from all participating mothers.

1.2 ***ALSPAC cohort description***

All pregnant women living in Avon (UK) between 1991 and 1992 were invited to take part in the ALSPAC study^3-5^. Of the total 20,248 eligible pregnancies identified, 14,541 were initially enrolled. Of the initial pregnancies, there was a total of 14,676 fetuses, resulting in 14,062 live births and 13,988 children were alive at 1 year of age. When children were around 7 years old, an attempt was made to bolster the initial sample with eligible cases who had failed to join the study originally. The total sample size for analyses using any data collected after the age of 7 is therefore 15,447 pregnancies, resulting in 15,658 fetuses. Of these 14,541 were live born singletons.

Gestational age (in days) and birth weight (in grams) were retrieved from medical records. At approximate ages 2, 9, 19, and 44 months, length/height and weight were collected in routine measurements performed by health visitors as part of the child health surveillance program and were extracted from the local child health database for the whole cohort. Around 10% of the whole cohort (children born in last 6 months of the recruitment phase) also participated in a sub-study was conducted where length/height and weight were measured on 10 occasions during research clinic visits at around ages 4, 8, 12, 18, 25, 31, 37, 43, 49, and 61 months. The whole cohort were also invited to attend research clinics at around ages 7, 8, 9, and 10 years where height and weight were measured by trained staff. Of the 14,541 singleton live births, 13,634 had data on gestational age at birth and birth weight, and of these, 13,580 had data on birth weight centile. After cleaning growth data and excluding offspring with fewer than two growth measurements between age 1 week and 10 years, there was a total of 12,899 offspring left for growth curve modelling.

Ethical approval for ALSPAC was obtained from the ALSPAC Law and Ethics committee and local research ethics committees. Informed consent for use of questionnaire and clinic data was obtained from participants following the recommendations of the ALSPAC Ethics and Law Committee at the time.

1.3 ***BiB cohort description***

All pregnant women living in Bradford (UK) between 2007 and 2011 were invited to participate in BiB during their oral glucose tolerance test, which is offered to all women booked for delivery at Bradford Royal Infirmary^6^. Of the 15,000 pregnant women invited, 12,453 women who experienced 13,776 pregnancies were enrolled into the study. BiB has almost an equal split of White European and South Asian women, all residing in Bradford, UK, a city in the North of England with high levels of socioeconomic deprivation. The BiB study was started due to a high prevalence of poor child health in the city. For the current study, only the first enrolled pregnancies were included.

Gestational age (in days) and birth weight (in grams) were retrieved from medical records^7^. Lngth/height, and weight measures from birth to age 10 years were performed by health visitors as part of standard care in the UK and were extracted from the child health records for the majority (98%) of the cohort. Length/height, and weight measurements up to age 10 years were also available from primary care records for some of the cohort. Around 1,700 families also participated in the BiB1000 sub-study where repeated measurements of height and weight were collected at home or in research clinics at ages 6, 12, 18, 24, and 36 months. Around age 4–5 years, height and weight measurements were collected by school nurses as part of the National Child Measurement Programme. The whole BiB cohort were also invited to attend a research clinic at age 7-10 years, where height and weight were measured by trained staff^8^. Of the 12,356 singleton live births, 12,057 had data on gestational age at birth and birth weight, and of these, 12,040 had data on birth weight centile. After cleaning growth data and excluding offspring with fewer than two growth measurements between age 1 week and 10 years, there was a total of 11,436 offspring left for growth curve modelling.

BiB had ethical approval from Bradford Research Ethics Committee (07/H1302/112). All BiB participants provided informed consent or assent to participate in the study and secondary data analyses.

1.4 ***EDEN cohort description***

All pregnant women attending their prenatal visit before 24 weeks gestation at Nancy and Poitiers university hospitals in Paris (France) between 2003 and 2006 were invited to participate in EDEN^9^. Exclusion criteria were multiple pregnancies, diabetes before pregnancy, French illiteracy, or any plans to move out of the region in the next 3 years. Of 3,758 women that were invited to participate, 2,002 were enrolled in the study.

Gestational age (in days) and birth weight (in grams) were retrieved from medical records. length/height, and weight measures from birth to age 10 years were performed by health professionals and retrieved from the child health booklet by parent reporting. Height and weight were also measured during research clinic visits at ages 1, 3, and 5. Of the 2,002 singleton live births, 1,899 had data on gestational age at birth and birth weight, and of these, 1,899 had data on birth weight centile. After cleaning growth data and excluding offspring with fewer than two growth measurements between age 1 week and 10 years, there was a total of 1,744 offspring left for growth curve modelling.

The EDEN study received approval from the ethics committee (CCPPRB) of Kremlin Bicêtre on 12 December 2002 and from CNIL (Commission Nationale Informatique et Liberté), the French data privacy institution. All participants gave their informed consent for inclusion before they participated in the study. Consent for the child was obtained from both parents after birth.

1.5 ***Generation XXI cohort description***

All pregnant women delivering live-born infants with gestational age above 24 weeks at all five public maternity units in the metropolitan area of Porto (Portugal) between 2005 and 2006, were invited to participate in Generation XXI ^10^. Of the 9,234 women that were invited, 8,495 women were enrolled and gave birth to 8,647 babies.

Gestational age (in days) and birth weight (in grams) were retrieved from obstetric records in Generation XXI ^7^. Length/height, and weight measures from birth to age 10 years were performed by health professionals as part of standard childcare in Portugal and were abstracted from the Child Health Book. Height and weight were also measured during research clinic visits at ages 4,7, and 10 years. Of the 8,351 singleton live births, 8,099 had data on gestational age at birth and birth weight, and of these, 8,093 had data on birth weight centile. After cleaning growth data and excluding offspring with fewer than two growth measurements between age 1 week and 10 years, there was a total of 6,377 offspring left for growth curve modelling.

The University of Porto Medical School/S. Joao Hospital Centre Ethics Committee approved the study protocols. Generation XXI complies with the Ethical Principles expressed in the Helsinki Declaration and with the national legislation and was registered with the Portuguese Authority for Data Protection. In all evaluations, Generation XXI participants were informed about the purposes and design of the study, as well as the potential discomfort caused by participation. Signed informed consent was obtained from all parents or legal guardians, and oral assent was obtained from children at each evaluation.

2. ***Replication cohort***
   2.1 ***Project Viva cohort description***

Pregnant women attending their 1^st^ trimester antenatal ultrasound scan at 8 obstetric offices of a multi-site group practice (Atrius Harvard Vanguard Medical Associations) in Eastern Massachusetts (USA), between 1999 and 2002 were invited to participate in Project Viva^11^. Exclusion criteria included multiple gestation, inability to answer questions in English, gestational age⩾22 weeks at recruitment, and plans to move from the study area before delivery. Of the 2,341 eligible women that were invited, 2,218 women and babies were enrolled.

Gestational age (in days) and birth weight (in grams) were retrieved from medical records. Length/height, and weight measurements from birth to age 10 years were obtained from medical records where paediatric clinics recorded length/height and weight at routine well-child visits during infancy and childhood. Height and weight were also measured by trained research assistants using standardized protocols during research clinics at mean age 6 months; 3 years, and 7 years. Of the 2,218 singleton live births, 2,127 had data on gestational age at birth and birth weight, and of these, 2,126 had data on birth weight centile. After cleaning growth data and excluding offspring with fewer than two growth measurements between age 1 week and 10 years, there was a total of 1,736 offspring left for growth curve modelling.

Mothers provided written informed consent at enrolment and each postnatal follow-up visit. The institutional review board of Harvard Pilgrim Health Care approved this project in line with ethical standards established by Declaration of Helsinki.

2.2 ***GUSTO cohort description***

All pregnant women attending their 1^st^ trimester antenatal ultrasound scan at Singapore’s two major public maternity units between 2009 and 2010 were invited to participate in GUSTO^10^. Women were eligible if they were aged 18 years or older, Singaporean citizens or permanent residents, with self-reported homogenous ethnic ancestry (Chinese, Indian, Malay), intended to deliver at either of the recruitment hospitals and reside in Singapore for the next five years. Women >14 weeks gestation, receiving chemotherapy, psychotropic medications, or having an existing type I diabetes mellitus diagnosis at time of recruitment were excluded. Women who ultimately did not agree to donate birth tissues (cord, placenta, cord blood) were also excluded. Of the 2,034 eligible women that were invited, 1,177 women were enrolled and gave birth to 1,176 babies.

Gestational age (in days) and birth weight (in grams) were retrieved from medical records^7^. Length/height, and weight measurements from birth to age 10 years were obtained during research clinic visits at around ages 3, 6, 9, 12, 15, 18, 24, 36 and 48 months, and ages 4.5, 5, 5.5, 6, 6.5, 7, 8, 9, and 10 years. Of the 1,176 singleton live births, 1,095 offspring had data on gestational age at birth and birth weight, and of these, 1,094 had data on birth weight centile. After cleaning growth data and excluding offspring with fewer than two growth measurements between age 1 week and 10 years, there was a total of 1,013 offspring left for growth curve modelling.

GUSTO study protocols following the principles of the Declaration of Helsinki and were approved by the respective ethics committees for two hospitals: National Healthcare Group Domain Specific Review Board (NUH) and SingHealth Centralized Institutional Review Board (KKH). All participants in this study provided informed consent to participate and contribute their data to publications. The GUSTO study is registered under study ID: NCT01174875 (clinicaltrials.gov) which broadly covers investigations of parental and gestational influences on child health.

3. ***Growth curve modelling and estimation of growth features***

Growth curve models for estimation of infant/child growth features were fitted separately in the Discovery (pooled ABCD, ALSPAC, BiB, EDEN, Generation XXI cohorts) and Replication cohorts (pooled Project Viva and GUSTO cohorts), and separately by sex. The analysis samples for growth curve modelling were all children with data on birth weight centile and two or more repeated measures between age 1 week and 10 years (n=38,342 in the Discovery cohort and n=2,749 in Replication cohort). The median (interquartile range) number of measurements per child was 7 (8) in the Discovery cohort and 14 (8) in the Replication cohort.

Individual weight, height, and BMI trajectories from age 1 week to 10 years were modelled using P-spline linear mixed effects models^12,13^. Each model included population-level (fixed effects) and individual-specific (random effects) nonlinear age functions estimated via cubic P-splines^14,15^. Models were fitted separately in boys and girls due to expected sex-differences in growth trajectories. P-splines were constructed using 10 evenly spaced cubic B-spline basis functions, with second-order difference penalties applied to the fixed effects and first-order difference penalties applied to the random effects basis. A square root transformation was applied to age to improve fit. Models and smoothing parameters were estimated using restricted maximum likelihood (REML). Models were fitted using the psme package in R^16^. The predicted growth trajectories were estimated by evaluating the fitted B-splines on an age grid of 550 values (equivalent to one-week intervals from age 1 week to 10 years) and using the estimated coefficients to compute the fixed (population-level) and random (individual-specific) predictions at each age. Predicted height, weight, and BMI trajectories from the fitted models were used to estimate growth outcomes for subsequent analysis.

Growth rate during infancy was estimated by calculating the first derivatives of the fitted height and weight trajectories and identifying the growth velocity at ages 1, 6, 12, and 24 months. To examine height and weight Z-score trajectories, we used our fitted trajectories to predict height and weight values at 3-month intervals from age 1-60 months and converted these to height-for-age and weight-for-age Z-scores using the WHO Child Growth Standards. Calculation of Z-scores was done using the zscorer package in R. The predicted BMIs up to, and after, age 2 years were used to calculate BMI and age at infant BMI peak and child BMI rebound, respectively. To estimate BMI at peak and rebound, we identified points where the second derivative changed sign: change from positive to negative indicates a peak and change from negative to positive indicates a trough (i.e., rebound). We used a quadratic fit around the turning points to improve accuracy of peak/trough location. A quadratic curve was fitted to a subset of data around the peak/trough location, and this subset was increased incrementally until a well-defined quadratic curve was found. Age at BMI peak and rebound was calculated using the formula for the vertex of a parabola. Corresponding BMIs at the peak/rebound age were predicted from the quadratic model. Calculation of BMI and age at peak/rebound BMI was done using the getPeakTrough() function from the sitar library^17^. Lastly, we applied the International Obesity Task Force age- and sex-specific age and sex-specific cut off points^18^ on our predicted BMIs at age 10 years to calculate the prevalence of overweight or obesity.

Children for whom we did not identify a BMI peak and/or rebound were excluded, which left a total of 36,018 and 2,517 children from the Discovery and Replication cohort respectively for subsequent association analyses. Pearson correlation matrices between continuous growth outcomes (i.e., height and weight velocity, age and BMI at peak BMI and rebound BMI) were calculated to aid in the interpretation of the subsequent association results.

4. ***Association analyses***
   4.1 ***Main analysis and replication***

All main association analyses were done separately in the Discovery and Replication cohorts, and in boys and girls combined. Mean differences in infant growth velocity (at ages 1, 6, 12, and 24 months) and BMI and age at infant peak BMI and childhood rebound BMI for each birth weight group (vs. first decile) were estimated using linear regression. Mean height-for-age and weight-for-age Z-score trajectory for each birth weight decile group from 1 month to 5 years were estimated using linear mixed effects models with random intercepts for child. Logistic regression to estimate mean differences in probability of overweight or obesity at age 10 years for each birth weight group (vs. first decile). For the latter, we analysed birth weight as a 10-level integer variable (i.e., per category change) in the Replication cohort due to low cell numbers in this cohort and this analysis was also done in the Discovery cohort to obtain the comparable estimate. All models were adjusted for sex and birth cohort. Z-scores trajectory models included a natural spline for age (as a fixed effect)^19^ to allow for nonlinear change in height and weight Z-score trajectories plus its interaction with birth weight decile group.

Results were presented as the predicted means (for Z-score analysis) or differences in predicted means (for other continuous growth outcomes) and predicted mean probability for overweight or obesity at age 10 years (and 95% confidence intervals) by birth weight decile group. Meta-regression was used to assess statistical evidence of replication by quantifying the difference in estimates between the Discovery and Replication cohorts. Separate random effects meta-analysis models were fitted for each outcome, using 'cohort' as a dichotomous moderator (i.e., Discovery vs. Replication cohort) and calculating the P-value for cohort. Meta-regression analysis was done using the metafor^20^ package in R.

4.2 ***Additional analysis in the Discovery cohort***

Additional analyses were conducted in the Discovery cohort. Sex differences were examined by fitting models with interactions between birth weight decile and sex and calculating the P-value for the interaction term. To evaluate whether using birth weight decile groups offered a predictive benefit over the three conventional groups (SGA, AGA, LGA), we used coefficient of determination (R^2^) and area under the receiver operating characteristics curve to compare the performance of two competing linear and logistic models, respectively (i.e., models with birth weight entered as decile groups vs. models with three (SGA, AGA, LGA) groups. We interrogated the nonlinear nature of associations by comparing models with birth weight (Z-score) as a linear term and as natural spline terms with 2-5 knots, using the BIC to select the best fitting model. The predicted mean across the entire birth weight Z-score range was then plotted for each model that showed evidence for nonlinear associations.

Lastly, we explored robustness of our results to measured confounders (factors that could influence both pre and postnatal growth) by fitting models with adjustment for pregnancy-related covariates (maternal parity, maternal ethnicity, and both parents’ BMI, age, smoking, and education). We used Directed Acyclic Graphs (DAG) to illustrate the relations between confounders, birth weight, and infant/child outcomes and selected the following maternal and paternal pregnancy-related factors measured in all cohorts for adjustment (in addition to sex and cohort adjustment): maternal parity, maternal ethnicity or birth region, and both parents’ BMI, age, smoking, and education. Parental pregnancy-related variables were measured by maternal self-report in questionnaires administered during pregnancy (at delivery in the Generation XXI cohort) or from medical records in all cohorts. For our analysis, we used variables that were harmonised using the lowest common denominator approach as part of the EU Child Cohort Network (ECCN)^21,22^. Maternal and paternal education were coded as low, medium, or high. Because some ECCN cohorts (e.g., EDEN cohort) were prohibited by law from collecting data on participants’ ethnicity, maternal ethnicity was derived using the best proxy in each study. For cohorts that recorded country of origin (based on maternal grandmother’s country of birth), mothers were classified as Western or Non‑Western. The Western group included the EU, Andorra, Australia, Canada, Iceland, Monaco, Liechtenstein, New Zealand, Norway, San Marino, Switzerland, United States, and Vatican City; all other countries were classified as Non‑Western. For cohorts that captured self‑reported skin colour, these were categorised as White, non‑White, or Mixed. In each cohort, whichever proxy (country of origin or skin colour) was deemed most reliable, was used to classify maternal ethnicity as Western versus Non‑Western/Mixed. Continuous measures of maternal and paternal age, and maternal and paternal BMI, and dichotomous measures of maternal parity (primiparous vs. multiparous), maternal and paternal smoking (no vs. yes) were derived.

Robustness to confounders was examined by refitting our main models with extra adjustment for measured pregnancy-related covariates. Missing data on the covariates was handled using multiple imputation by chained equations, under the assumption that, given the variables in the imputation procedure, the missing data were missing at random. There were more missing values for paternal than maternal covariates: proportion of missing values ranged from 3.8% (parity) to 13.3% (BMI) in mothers and 32.2% (education) to 54.7% (smoking) in fathers. A total of 20 imputed datasets were generated. Imputation models included cohort, sex, birth weight, gestational age, birth weight deciles, birth weight Z-score, and all growth outcomes and covariates (maternal parity, both parents’ education, BMI, smoking, and age). We used Rubin's Rules to combine estimates across imputed datasets. Multiple imputation was done using the mice package in R^23^. Fully adjusted analyses were compared with analyses adjusted for only the maternal factors: results from both were similar and so only the fully adjusted results are presented. Lastly, we assessed our imputation model by comparing with analyses restricted to complete cases (adjusting for maternal covariates).

All analysis was done using R 4.5.2 (R Project for Statistical Computing).

***References***

13.	María Alejandra Hernandez ZL, Tim J Cole, Yi Ong, Kate Tilling, Ahmed Elhakeem. Capturing infant and child growth dynamics with P-splines mixed effects models. *medRxiv*. 2025;

14.	Eilers PHC, Marx BD. Flexible smoothing with B-splines and penalties. *Statistical Science*. 1996;11(2):89-121, 33.

15.	Eilers PHC, Marx BD. *Practical Smoothing: The Joys of P-splines*. Cambridge University Press; 2021.

### eTable 1 Birth and infant/child growth outcomes in each individual birth cohort

|  | ABCD,  n=5678 | ALSPAC,  n=12277 | BiB,  n=10512 | EDEN,  n=1698 | Generation XXI,  n=5853 | Project Viva, n=1607 | GUSTO, n=910 |
| --- | --- | --- | --- | --- | --- | --- | --- |
| Sex, No. (%) |  |  |  |  |  |  |  |
| *Female* | 2821 (49.7) | 5866 (47.8) | 4978 (47.4) | 806 (47.5) | 2767 (47.3) | 765 (47.6) | 426 (46.8) |
| *Male* | 2857 (50.3) | 6411 (52.2) | 5534 (52.6) | 892 (52.5) | 3086 (52.7) | 842 (52.4) | 484 (53.2) |
| Birth weight, mean (SD), g | 3453 (549) | 3407 (542) | 3223 (548) | 3288 (504) | 3192 (486) | 3469 (582) | 3104 (443) |
| Gestational age, mean (SD), wk | 39.8 (1.7) | 40.0 (1.8) | 39.5 (1.8) | 39.7 (1.7) | 39.4 (1.6) | 39.4 (1.9) | 38.8 (1.4) |
| Birth weight centile, mean (SD), % | 61.1 (28.3) | 56.8 (29.3) | 47.8 (30.0) | 51.1 (28.4) | 47.3 (27.6) | 65.1 (27.5) | 45.8 (27.1) |
| Birth weight centile groups, No. (%) |  |  |  |  |  |  |  |
| *1^st^* | 302 (5.3) | 1007 (8.2) | 1367 (13.0) | 146 (8.6) | 614 (10.5) | 53 (3.3) | 92 (10.1) |
| *2^nd^* | 353 (6.2) | 888 (7.2) | 1177 (11.2) | 169 (10.0) | 619 (10.6) | 80 (5.0) | 113 (12.4) |
| *3^rd^* | 382 (6.7) | 935 (7.6) | 1045 (9.9) | 173 (10.2) | 630 (10.8) | 114 (7.1) | 97 (10.7) |
| *4^th^* | 400 (7.0) | 984 (8.0) | 985 (9.4) | 161 (9.5) | 658 (11.2) | 99 (6.2) | 115 (12.6) |
| *5^th^* | 527 (9.3) | 1091 (8.9) | 1026 (9.8) | 172 (10.1) | 645 (11.0) | 127 (7.9) | 90 (9.9) |
| *6^th^* | 534 (9.4) | 1185 (9.7) | 943 (9.0) | 178 (10.5) | 632 (10.8) | 138 (8.6) | 106 (11.6) |
| *7^th^* | 597 (10.5) | 1292 (10.5) | 991 (9.4) | 178 (10.5) | 571 (9.8) | 168 (10.5) | 95 (10.4) |
| *8^th^* | 706 (12.4) | 1419 (11.6) | 881 (8.4) | 179 (10.5) | 536 (9.2) | 196 (12.2) | 70 (7.7) |
| *9^th^* | 800 (14.1) | 1528 (12.4) | 1006 (9.6) | 181 (10.7) | 550 (9.4) | 248 (15.4) | 75 (8.2) |
| *10^th^* | 1077 (19.0) | 1948 (15.9) | 1091 (10.4) | 161 (9.5) | 398 (6.8) | 384 (23.9) | 57 (6.3) |
| Height velocity, mean (SD), cm/mo |  |  |  |  |  |  |  |
| *1 mo* | 2.3 (0.2) | 2.3 (0.1) | 2.3 (0.2) | 2.3 (0.2) | 2.4 (0.2) | 2.3 (0.2) | 2.4 (0.2) |
| *6 mo* | 2.4 (0.2) | 2.3 (0.2) | 2.4 (0.2) | 2.4 (0.2) | 2.5 (0.3) | 2.3 (0.2) | 2.4 (0.2) |
| *12 mo* | 2.4 (0.2) | 2.3 (0.2) | 2.3 (0.2) | 2.4 (0.2) | 2.4 (0.2) | 2.4 (0.2) | 2.4 (0.2) |
| *24 mo* | 2.4 (0.2) | 2.3 (0.2) | 2.3 (0.2) | 2.4 (0.2) | 2.4 (0.2) | 2.3 (0.2) | 2.2 (0.2) |
| Weight velocity, mean (SD), g/mo |  |  |  |  |  |  |  |
| *1 mo* | 577 (123) | 597 (89) | 598 (101) | 584 (122) | 604 (125) | 679 (136) | 697 (140) |
| *6 mo* | 647 (120) | 658 (106) | 655 (106) | 653 (127) | 680 (131) | 611 (126) | 545 (116) |
| *12 mo* | 558 (114) | 551 (108) | 553 (105) | 543 (105) | 584 (119) | 541 (114) | 462 (112) |
| *24 mo* | 558 (123) | 536 (126) | 542 (121) | 532 (116) | 567 (135) | 581 (128) | 528 (135) |
| Peak BMI, mean (SD), kg/m^2^ | 17.4 (1.2) | 17.6 (1.2) | 17.3 (1.2) | 17.4 (1.2) | 17.7 (1.3) | 17.9 (1.3) | 17.2 (1.2) |
| Age at peak BMI, mean (SD), mo | 9.6 (2.6) | 10.5 (2.5) | 10.4 (2.4) | 9.5 (2.5) | 10.0 (2.9) | 7.9 (2.5) | 6.6 (1.9) |
| Rebound BMI, mean (SD), kg/m^2^ | 15.3 (1.1) | 15.6 (1.2) | 15.5 (1.3) | 15.3 (1.0) | 15.7 (1.3) | 15.8 (1.2) | 14.9 (1.3) |
| Age at rebound BMI, mean (SD), y | 5.2 (1.3) | 5.3 (1.5) | 5.3 (1.6) | 5.0 (1.4) | 4.8 (1.6) | 5.0 (1.5) | 5.2 (1.6) |
| Overweight or obesity at 10 y, No. (%) |  |  |  |  |  |  |  |
| *No* | 5388 (94.9) | 10973 (89.4) | 9182 (87.3) | 1615 (95.1) | 4553 (77.8) | 1327 (82.6) | 772 (84.8) |
| *Yes* | 290 (5.1) | 1304 (10.6) | 1330 (12.7) | 83 (4.9) | 1300 (22.2) | 280 (17.4) | 138 (15.2) |

### eTable 2. Sex and cohort-adjusted mean differences in infant/child growth outcomes in the Discovery and Replication cohorts

| Birth weight decile | Discovery cohort | Replication cohort | *P* cohort difference |
| --- | --- | --- | --- |
|  | Height velocity at age 1 mo, cm/mo | | 0.03 |
| *1^st^* | ref | ref |  |
| *2^nd^* | -0.02 (-0.03 to -0.01) | -0.03 (-0.07 to 0.01) |  |
| *3^rd^* | -0.03 (-0.04 to -0.02) | -0.04 (-0.08 to 0.00) |  |
| *4^th^* | -0.04 (-0.05 to -0.04) | -0.06 (-0.1 to -0.02) |  |
| *5^th^* | -0.04 (-0.05 to -0.04) | -0.07 (-0.12 to -0.03) |  |
| *6^th^* | -0.05 (-0.05 to -0.04) | -0.07 (-0.11 to -0.03) |  |
| *7^th^* | -0.05 (-0.06 to -0.04) | -0.05 (-0.09 to -0.01) |  |
| *8^th^* | -0.06 (-0.07 to -0.06) | -0.09 (-0.13 to -0.05) |  |
| *9^th^* | -0.07 (-0.07 to -0.06) | -0.1 (-0.14 to -0.06) |  |
| *10^th^* | -0.08 (-0.09 to -0.07) | -0.12 (-0.16 to -0.08) |  |
|  | Height velocity at age 6 mo, cm/mo | | 0.9 |
| *1^st^* | ref | ref |  |
| *2^nd^* | -0.03 (-0.04 to -0.02) | -0.01 (-0.06 to 0.03) |  |
| *3^rd^* | -0.04 (-0.05 to -0.03) | -0.02 (-0.07 to 0.03) |  |
| *4^th^* | -0.05 (-0.06 to -0.04) | -0.05 (-0.1 to -0.01) |  |
| *5^th^* | -0.06 (-0.07 to -0.04) | -0.06 (-0.11 to -0.01) |  |
| *6^th^* | -0.06 (-0.07 to -0.05) | -0.05 (-0.1 to 0.00) |  |
| *7^th^* | -0.06 (-0.07 to -0.05) | -0.07 (-0.11 to -0.02) |  |
| *8^th^* | -0.08 (-0.09 to -0.07) | -0.06 (-0.11 to -0.02) |  |
| *9^th^* | -0.08 (-0.09 to -0.07) | -0.1 (-0.15 to -0.06) |  |
| *10^th^* | -0.10 (-0.11 to -0.09) | -0.11 (-0.16 to -0.07) |  |
|  | Height velocity at age 12 mo, cm/mo | | 0.00002 |
| *1^st^* | ref | ref |  |
| *2^nd^* | -0.02 (-0.03 to -0.01) | 0.03 (-0.02 to 0.08) |  |
| *3^rd^* | -0.02 (-0.03 to -0.01) | 0.03 (-0.02 to 0.07) |  |
| *4^th^* | -0.02 (-0.03 to -0.01) | 0.0 (-0.04 to 0.05) |  |
| *5^th^* | -0.03 (-0.04 to -0.02) | 0.01 (-0.04 to 0.05) |  |
| *6^th^* | -0.02 (-0.03 to -0.01) | 0.02 (-0.02 to 0.06) |  |
| *7^th^* | -0.02 (-0.03 to -0.01) | 0 (-0.04 to 0.04) |  |
| *8^th^* | -0.03 (-0.04 to -0.02) | 0.02 (-0.02 to 0.06) |  |
| *9^th^* | -0.03 (-0.04 to -0.02) | -0.01 (-0.05 to 0.03) |  |
| *10^th^* | -0.04 (-0.05 to -0.03) | -0.02 (-0.06 to 0.02) |  |
|  | Height velocity at age 24 mo, cm/mo | | 0.002 |
| *1^st^* | ref | ref |  |
| *2^nd^* | 0.0 (-0.01 to 0.01) | 0.04 (-0.01 to 0.08) |  |
| *3^rd^* | 0.0 (-0.01 to 0.02) | 0.04 (0.0 to 0.09) |  |
| *4^th^* | 0.02 (0.01 to 0.03) | 0.03 (-0.02 to 0.07) |  |
| *5^th^* | 0.02 (0.01 to 0.03) | 0.05 (0 to 0.09) |  |
| *6^th^* | 0.02 (0.01 to 0.03) | 0.05 (0 to 0.09) |  |
| *7^th^* | 0.03 (0.02 to 0.04) | 0.06 (0.02 to 0.11) |  |
| *8^th^* | 0.03 (0.02 to 0.04) | 0.08 (0.04 to 0.12) |  |
| *9^th^* | 0.04 (0.03 to 0.05) | 0.06 (0.02 to 0.11) |  |
| *10^th^* | 0.05 (0.04 to 0.06) | 0.07 (0.03 to 0.11) |  |
|  | Weight velocity at age 1 mo, g/mo | | 0.06 |
| *1^st^* | ref | ref |  |
| *2^nd^* | 0.92 (-3.42 to 5.3) | 7.2 (-19.8 to 34.3) |  |
| *3^rd^* | 5.3 (0.93 to 9.6) | 10.9 (-15.7 to 37.5) |  |
| *4^th^* | 2.8 (-1.57 to 7.1) | 12.6 (-13.9 to 39.1) |  |
| *5^th^* | 4.7 (0.44 to 9.0) | 13.5 (-13.0 to 40.0) |  |
| *6^th^* | 5.6 (1.4 to 9.9) | 9.5 (-16.3 to 35.4) |  |
| *7^th^* | 5.6 (1.4 to 9.8) | 31.7 (6.1 to 57.3) |  |
| *8^th^* | 4.3 (0.07 to 8.5) | 8.7 (-17.0 to 34.4) |  |
| *9^th^* | 4.7 (0.62 to 8.8) | 24.6 (-0.33 to 49.6) |  |
| *10^th^* | 0.32 (-3.7 to 4.3) | -9.3 (-33.5 to 14.9) |  |
|  | Weight velocity at age 6 mo, g/mo | | 0.8 |
| *1^st^* | ref | ref |  |
| *2^nd^* | 1.4 (-4.0 to 6.8) | 6.32 (-20.0 to 32.7) |  |
| *3^rd^* | 9.1 (3.6 to 14.5) | 17.0 (-9.4 to 42.5) |  |
| *4^th^* | 9.8 (4.3 to 15.2) | 19.4 (-6.4 to 45.2) |  |
| *5^th^* | 10.7 (5.3 to 16.0) | 11.8 (-14.0 to 37.6) |  |
| *6^th^* | 14.7 (9.4 to 20.0) | 22.6 (-2.6 to 47.8) |  |
| *7^th^* | 18.6 (13.4 to 23.9) | 18.2 (-6.7 to 43.2) |  |
| *8^th^* | 21.9 (16.7 to 27.2) | 13.2 (-11.8 to 38.3) |  |
| *9^th^* | 22.1 (17.0 to 27.3) | 12.9 (-11.4 to 37.2) |  |
| *10^th^* | 24.6 (19.6 to 29.6) | 24.3 (0.70 to 47.8) |  |
|  | Weight velocity at age 12 mo, g/mo | | 0.2 |
| *1^st^* | ref | ref |  |
| *2^nd^* | 3.89 (-1.4 to 9.1) | 32.8 (8.4 to 57.1) |  |
| *3^rd^* | 10.6 (5.4 to 15.9) | 29.7 (5.7 to 53.7) |  |
| *4^th^* | 15.8 (10.6 to 21.1) | 20.6 (-3.3 to 44.4) |  |
| *5^th^* | 16.3 (11.2 to 21.5) | 19.7 (-4.2 to 43.6) |  |
| *6^th^* | 23.1 (17.9 to 28.2) | 41.8 (18.5 to 65.1) |  |
| *7^th^* | 29.9 (24.9 to 35.0) | 26.9 (3.8 to 50.0) |  |
| *8^th^* | 34.6 (29.6 to 39.7) | 35.3 (12.1 to 58.5) |  |
| *9^th^* | 37.2 (32.3 to 42.2) | 35.7 (13.2 to 58.2) |  |
| *10^th^* | 45.9 (41.1 to 50.7) | 54.3 (32.52 to 76.1) |  |
|  | Weight velocity at age 24 mo, g/mo | | 0.09 |
| *1^st^* | ref | ref |  |
| *2^nd^* | 8.12 (2.2 to 14.1) | 43.9 (16.0 to 71.7) |  |
| *3^rd^* | 16.2 (10.3 to 22.2) | 38.5 (11.1 to 65.9) |  |
| *4^th^* | 23.7 (17.7 to 29.6) | 24.0 (-3.3 to 51.3) |  |
| *5^th^* | 24.9 (19.1 to 30.7) | 45.8 (18.5 to 73.1) |  |
| *6^th^* | 33.6 (27.8 to 39.5) | 51.2 (24.5 to 77.8) |  |
| *7^th^* | 41.2 (35.4 to 46.9) | 56.0 (29.6 to 82.4) |  |
| *8^th^* | 47.3 (41.5 to 53.0) | 62.6 (36.2 to 89.1) |  |
| *9^th^* | 52.9 (47.3 to 58.6) | 63.9 (38.2 to 89.7) |  |
| *10^th^* | 68.9 (63.4 to 74.3) | 80.3 (55.4 to 105.2) |  |
|  | Infant peak BMI (kg/m^2^) | | 0.7 |
| *1^st^* | ref | ref |  |
| *2^nd^* | 0.23 (0.17 to 0.28) | 0.23 (-0.04 to 0.49) |  |
| *3^rd^* | 0.41 (0.36 to 0.47) | 0.41 (0.15 to 0.68) |  |
| *4^th^* | 0.49 (0.43 to 0.54) | 0.57 (0.31 to 0.83) |  |
| *5^th^* | 0.54 (0.49 to 0.6) | 0.66 (0.39 to 0.92) |  |
| *6^th^* | 0.68 (0.62 to 0.73) | 0.74 (0.48 to 0.99) |  |
| *7^th^* | 0.77 (0.72 to 0.82) | 0.83 (0.57 to 1.08) |  |
| *8^th^* | 0.84 (0.78 to 0.89) | 0.77 (0.52 to 1.03) |  |
| *9^th^* | 0.94 (0.89 to 0.99) | 1.08 (0.83 to 1.33) |  |
| *10^th^* | 1.16 (1.11 to 1.21) | 1.2 (0.96 to 1.44) |  |
|  | Age at infant peak BMI (mo) | | 0.001 |
| *1^st^* | ref | ref |  |
| *2^nd^* | -0.31 (-0.43 to -0.19) | 0.25 (-0.24 to 0.73) |  |
| *3^rd^* | -0.38 (-0.5 to -0.26) | 0.09 (-0.39 to 0.56) |  |
| *4^th^* | -0.4 (-0.52 to -0.28) | 0.11 (-0.37 to 0.58) |  |
| *5^th^* | -0.49 (-0.6 to -0.37) | -0.18 (-0.66 to 0.3) |  |
| *6^th^* | -0.58 (-0.7 to -0.46) | 0.04 (-0.43 to 0.5) |  |
| *7^th^* | -0.58 (-0.69 to -0.46) | -0.28 (-0.74 to 0.18) |  |
| *8^th^* | -0.52 (-0.64 to -0.41) | -0.36 (-0.83 to 0.1) |  |
| *9^th^* | -0.74 (-0.86 to -0.63) | -0.47 (-0.91 to -0.02) |  |
| *10^th^* | -0.96 (-1.07 to -0.85) | -0.48 (-0.92 to -0.05) |  |
|  | Childhood rebound BMI (kg/m^2^) | | 0.8 |
| *1^st^* | ref | ref |  |
| *2^nd^* | 0.17 (0.12 to 0.23) | 0.3 (0.04 to 0.55) |  |
| *3^rd^* | 0.32 (0.26 to 0.38) | 0.4 (0.15 to 0.65) |  |
| *4^th^* | 0.37 (0.31 to 0.43) | 0.38 (0.13 to 0.63) |  |
| *5^th^* | 0.4 (0.34 to 0.45) | 0.53 (0.28 to 0.78) |  |
| *6^th^* | 0.53 (0.47 to 0.59) | 0.58 (0.33 to 0.82) |  |
| *7^th^* | 0.62 (0.57 to 0.68) | 0.63 (0.39 to 0.87) |  |
| *8^th^* | 0.67 (0.61 to 0.72) | 0.58 (0.33 to 0.82) |  |
| *9^th^* | 0.74 (0.69 to 0.8) | 0.72 (0.48 to 0.95) |  |
| *10^th^* | 0.95 (0.89 to 1.0) | 0.93 (0.71 to 1.16) |  |
|  | Age at childhood rebound BMI (mo) | | 0.0008 |
| *1^st^* | ref | ref |  |
| *2^nd^* | -0.19 (-1.05 to 0.67) | -0.97 (-4.91 to 3.0) |  |
| *3^rd^* | -0.05 (-0.92 to 0.81) | -2.8 (-6.66 to 1.1) |  |
| *4^th^* | 0.1 (-0.77 to 0.96) | -2.2 (-6.0 to 1.7) |  |
| *5^th^* | 0.65 (-0.2 to 1.5) | -2.8 (-6.6 to 1.1) |  |
| *6^th^* | 0.25 (-0.59 to 1.1) | -2.58 (-6.3 to 1.2) |  |
| *7^th^* | 0.2 (-0.64 to 1.03) | -3.62 (-7.3 to 0.1) |  |
| *8^th^* | 0.91 (0.07 to 1.74) | -2.33 (-6.1 to 1.4) |  |
| *9^th^* | 0.85 (0.03 to 1.66) | -0.12 (-3.8 to 3.5) |  |
| *10^th^* | 0.19 (-0.6 to 0.99) | 0.09 (-3.4 to 3.6) |  |
|  | Overweight or obesity at age 10 years (%) | |  |
| *1^st^* | ref | - |  |
| *2^nd^* | 3 (-13 to 19) | - |  |
| *3^rd^* | 14 (-2 to 30) | - |  |
| *4^th^* | 15 (-1 to 31) | - |  |
| *5^th^* | 18 (2 to 33) | - |  |
| *6^th^* | 23 (7 to 38) | - |  |
| *7^th^* | 34 (18 to 49) | - |  |
| *8^th^* | 45 (30 to 60) | - |  |
| *9^th^* | 46 (31 to 61) | - |  |
| *10^th^* | 71 (57 to 86) | - |  |
| Per category change | 7 (6 to 9) | 9 (5 to 13) | 0.6 |

### eTable 3. Parental pregnancy-related factors in the Discovery cohort

| Maternal pre-pregnancy BMI (kg/m^2^), mean (SD) | 23.9 (4.6) |
| --- | --- |
| Paternal BMI (kg/m^2^), mean (SD) | 25.4 (3.8) |
| Maternal age at pregnancy (years), mean (SD) | 28.4 (5.7) |
| Paternal age (years), mean (SD) | 31.9 (6.1) |
| Maternal parity: |  |
| *Primiparous, n (%)* | 19182 (55.4) |
| *Multiparous, n (%)* | 15473 (44.6) |
| Maternal early pregnancy smoking: |  |
| *No, n (%)* | 25788 (78.1) |
| *Yes, n (%)* | 7231 (21.9) |
| Paternal smoking: |  |
| *No, n (%)* | 10536 (64.6) |
| *Yes, n (%)* | 5763 (35.4) |
| Maternal education: |  |
| *High, n (%)* | 8041 (24.9) |
| *Medium, n (%)* | 13881 (43.1) |
| *Low, n (%)* | 10268 (31.9) |
| Paternal education: |  |
| *High, n (%)* | 6987 (28.9) |
| *Medium, n (%)* | 8949 (37.0) |
| *Low, n (%)* | 8241 (34.1) |
| Maternal ethnicity/birth region: |  |
| *Western, n (%)* | 25314 (78.3) |
| *Non-Western/Mixed, n (%)* | 7019 (21.7) |

### eTable 4. Confounder-adjusted mean differences in infant/child growth outcomes in the Discovery cohort

| Birth weight decile | Multiple imputation analysis, n=36,018 | Complete-case-analysis, n=27,500 |
| --- | --- | --- |
|  | Height velocity at age 1 mo, cm/mo | |
| *1^st^* | ref | ref |
| *2^nd^* | -0.02 (-0.03 to -0.01) | -0.03 (-0.03 to -0.02) |
| *3^rd^* | -0.03 (-0.03 to -0.02) | -0.03 (-0.04 to -0.02) |
| *4^th^* | -0.04 (-0.05 to -0.03) | -0.04 (-0.05 to -0.04) |
| *5^th^* | -0.04 (-0.05 to -0.03) | -0.04 (-0.05 to -0.04) |
| *6^th^* | -0.04 (-0.05 to -0.03) | -0.05 (-0.06 to -0.04) |
| *7^th^* | -0.04 (-0.05 to -0.04) | -0.05 (-0.06 to -0.04) |
| *8^th^* | -0.06 (-0.06 to -0.05) | -0.06 (-0.07 to -0.05) |
| *9^th^* | -0.06 (-0.07 to -0.05) | -0.07 (-0.08 to -0.06) |
| *10^th^* | -0.07 (-0.08 to -0.06) | -0.08 (-0.09 to -0.07) |
|  | Height velocity at age 6 mo, cm/mo | |
| *1^st^* | ref | ref |
| *2^nd^* | -0.03 (-0.04 to -0.01) | -0.04 (-0.05 to -0.02) |
| *3^rd^* | -0.03 (-0.04 to -0.02) | -0.04 (-0.05 to -0.03) |
| *4^th^* | -0.04 (-0.05 to -0.03) | -0.05 (-0.06 to -0.04) |
| *5^th^* | -0.05 (-0.06 to -0.04) | -0.06 (-0.07 to -0.04) |
| *6^th^* | -0.05 (-0.06 to -0.04) | -0.06 (-0.07 to -0.04) |
| *7^th^* | -0.05 (-0.06 to -0.04) | -0.06 (-0.07 to -0.05) |
| *8^th^* | -0.06 (-0.07 to -0.05) | -0.07 (-0.08 to -0.06) |
| *9^th^* | -0.07 (-0.08 to -0.06) | -0.08 (-0.09 to -0.06) |
| *10^th^* | -0.08 (-0.09 to -0.07) | -0.09 (-0.1 to -0.08) |
|  | Height velocity at age 12 mo, cm/mo | |
| *1^st^* | ref | ref |
| *2^nd^* | -0.01 (-0.02 to 0) | -0.02 (-0.03 to -0.01) |
| *3^rd^* | -0.02 (-0.03 to -0.01) | -0.02 (-0.03 to -0.01) |
| *4^th^* | -0.01 (-0.02 to 0) | -0.02 (-0.03 to 0) |
| *5^th^* | -0.02 (-0.03 to -0.01) | -0.03 (-0.04 to -0.01) |
| *6^th^* | -0.02 (-0.03 to -0.01) | -0.02 (-0.03 to -0.01) |
| *7^th^* | -0.02 (-0.03 to -0.01) | -0.02 (-0.03 to -0.01) |
| *8^th^* | -0.02 (-0.03 to -0.01) | -0.02 (-0.03 to -0.01) |
| *9^th^* | -0.02 (-0.03 to -0.01) | -0.03 (-0.04 to -0.01) |
| *10^th^* | -0.02 (-0.03 to -0.01) | -0.03 (-0.04 to -0.02) |
|  | Height velocity at age 24 mo, cm/mo | |
| *1^st^* | ref | ref |
| *2^nd^* | 0 (-0.01 to 0.01) | 0 (-0.01 to 0.02) |
| *3^rd^* | 0.01 (0 to 0.02) | 0.01 (-0.01 to 0.02) |
| *4^th^* | 0.02 (0.01 to 0.03) | 0.02 (0.01 to 0.03) |
| *5^th^* | 0.02 (0.01 to 0.03) | 0.02 (0.01 to 0.03) |
| *6^th^* | 0.02 (0.01 to 0.03) | 0.02 (0.01 to 0.03) |
| *7^th^* | 0.03 (0.02 to 0.04) | 0.03 (0.02 to 0.05) |
| *8^th^* | 0.04 (0.03 to 0.05) | 0.04 (0.03 to 0.05) |
| *9^th^* | 0.04 (0.03 to 0.05) | 0.04 (0.03 to 0.05) |
| *10^th^* | 0.05 (0.04 to 0.06) | 0.06 (0.05 to 0.07) |
|  | Weight velocity at age 1 mo, g/mo | |
| *1^st^* | ref | ref |
| *2^nd^* | 2.23 (-2.1 to 6.55) | -1.64 (-6.78 to 3.49) |
| *3^rd^* | 7.06 (2.71 to 11.4) | 4.37 (-0.79 to 9.52) |
| *4^th^* | 5.38 (1.04 to 9.72) | 2.86 (-2.29 to 8) |
| *5^th^* | 7.05 (2.8 to 11.31) | 5.88 (0.83 to 10.94) |
| *6^th^* | 8.68 (4.41 to 12.95) | 4.64 (-0.43 to 9.7) |
| *7^th^* | 8.34 (4.11 to 12.57) | 5.3 (0.27 to 10.33) |
| *8^th^* | 7.96 (3.73 to 12.19) | 4.8 (-0.19 to 9.8) |
| *9^th^* | 8.27 (4.11 to 12.43) | 5.4 (0.48 to 10.31) |
| *10^th^* | 4.54 (0.44 to 8.64) | 1.48 (-3.38 to 6.35) |
|  | Weight velocity at age 6 mo, g/mo | |
| *1^st^* | ref | ref |
| *2^nd^* | 2.96 (-2.43 to 8.35) | -1.47 (-7.77 to 4.83) |
| *3^rd^* | 11.19 (5.78 to 16.6) | 8.73 (2.4 to 15.06) |
| *4^th^* | 12.77 (7.36 to 18.18) | 10.07 (3.76 to 16.39) |
| *5^th^* | 13.47 (8.17 to 18.77) | 10.94 (4.74 to 17.14) |
| *6^th^* | 18.25 (12.93 to 23.57) | 13.35 (7.14 to 19.57) |
| *7^th^* | 21.91 (16.63 to 27.18) | 20.01 (13.83 to 26.18) |
| *8^th^* | 26.3 (21.02 to 31.57) | 24.34 (18.2 to 30.47) |
| *9^th^* | 26.23 (21.05 to 31.41) | 22.92 (16.89 to 28.95) |
| *10^th^* | 29.97 (24.87 to 35.08) | 27.92 (21.95 to 33.9) |
|  | Weight velocity at age 12 mo, g/mo | |
| *1^st^* | ref | ref |
| *2^nd^* | 5.04 (-0.18 to 10.26) | 2.45 (-3.61 to 8.52) |
| *3^rd^* | 12.28 (7.04 to 17.52) | 10.05 (3.96 to 16.14) |
| *4^th^* | 17.92 (12.67 to 23.17) | 16.6 (10.52 to 22.67) |
| *5^th^* | 18.15 (13.02 to 23.29) | 15.29 (9.32 to 21.25) |
| *6^th^* | 25.38 (20.23 to 30.53) | 23.49 (17.51 to 29.47) |
| *7^th^* | 32.25 (27.14 to 37.35) | 31.92 (25.97 to 37.86) |
| *8^th^* | 37.62 (32.51 to 42.73) | 37.71 (31.81 to 43.61) |
| *9^th^* | 39.87 (34.85 to 44.89) | 38.32 (32.51 to 44.12) |
| *10^th^* | 49.84 (44.89 to 54.78) | 49.85 (44.11 to 55.6) |
|  | Weight velocity at age 24 mo, g/mo | |
| *1^st^* | ref | ref |
| *2^nd^* | 9.1 (3.2 to 15.01) | 9.4 (2.53 to 16.27) |
| *3^rd^* | 17.43 (11.49 to 23.37) | 14.71 (7.81 to 21.6) |
| *4^th^* | 25.05 (19.1 to 30.99) | 25.27 (18.39 to 32.16) |
| *5^th^* | 25.73 (19.92 to 31.54) | 24.12 (17.36 to 30.88) |
| *6^th^* | 34.73 (28.9 to 40.56) | 35.58 (28.81 to 42.36) |
| *7^th^* | 41.95 (36.17 to 47.73) | 42.45 (35.72 to 49.18) |
| *8^th^* | 48.47 (42.68 to 54.25) | 47.07 (40.38 to 53.75) |
| *9^th^* | 53.36 (47.68 to 59.04) | 53.97 (47.4 to 60.54) |
| *10^th^* | 69.26 (63.66 to 74.86) | 69.46 (62.95 to 75.97) |
|  | Infant peak BMI (kg/m^2^) | |
| *1^st^* | ref | ref |
| *2^nd^* | 0.24 (0.18 to 0.29) | 0.21 (0.14 to 0.27) |
| *3^rd^* | 0.42 (0.37 to 0.48) | 0.39 (0.32 to 0.45) |
| *4^th^* | 0.5 (0.44 to 0.55) | 0.49 (0.43 to 0.56) |
| *5^th^* | 0.55 (0.49 to 0.6) | 0.53 (0.47 to 0.59) |
| *6^th^* | 0.68 (0.63 to 0.74) | 0.64 (0.58 to 0.7) |
| *7^th^* | 0.77 (0.71 to 0.82) | 0.74 (0.68 to 0.8) |
| *8^th^* | 0.84 (0.79 to 0.9) | 0.82 (0.76 to 0.89) |
| *9^th^* | 0.93 (0.88 to 0.99) | 0.91 (0.85 to 0.98) |
| *10^th^* | 1.14 (1.09 to 1.19) | 1.12 (1.06 to 1.19) |
|  | Age at infant peak BMI (mo) | |
| *1^st^* | ref | ref |
| *2^nd^* | -0.31 (-0.43 to -0.19) | -0.35 (-0.49 to -0.21) |
| *3^rd^* | -0.39 (-0.51 to -0.26) | -0.42 (-0.56 to -0.28) |
| *4^th^* | -0.41 (-0.53 to -0.28) | -0.42 (-0.56 to -0.28) |
| *5^th^* | -0.5 (-0.62 to -0.38) | -0.54 (-0.68 to -0.4) |
| *6^th^* | -0.6 (-0.72 to -0.48) | -0.66 (-0.8 to -0.52) |
| *7^th^* | -0.6 (-0.72 to -0.48) | -0.58 (-0.71 to -0.44) |
| *8^th^* | -0.55 (-0.67 to -0.43) | -0.55 (-0.69 to -0.41) |
| *9^th^* | -0.78 (-0.89 to -0.66) | -0.81 (-0.95 to -0.68) |
| *10^th^* | -1 (-1.12 to -0.89) | -1.01 (-1.14 to -0.87) |
|  | Childhood rebound BMI (kg/m^2^) | |
| *1^st^* | ref | ref |
| *2^nd^* | 0.19 (0.13 to 0.24) | 0.19 (0.13 to 0.26) |
| *3^rd^* | 0.33 (0.28 to 0.39) | 0.3 (0.23 to 0.36) |
| *4^th^* | 0.39 (0.33 to 0.44) | 0.4 (0.34 to 0.46) |
| *5^th^* | 0.4 (0.35 to 0.45) | 0.41 (0.34 to 0.47) |
| *6^th^* | 0.53 (0.48 to 0.59) | 0.52 (0.46 to 0.58) |
| *7^th^* | 0.61 (0.55 to 0.66) | 0.62 (0.56 to 0.68) |
| *8^th^* | 0.66 (0.61 to 0.72) | 0.64 (0.58 to 0.71) |
| *9^th^* | 0.72 (0.66 to 0.77) | 0.74 (0.68 to 0.8) |
| *10^th^* | 0.89 (0.84 to 0.94) | 0.9 (0.84 to 0.96) |
|  | Age at childhood rebound BMI (mo) | |
| *1^st^* | ref | ref |
| *2^nd^* | -0.43 (-1.28 to 0.42) | -0.95 (-1.93 to 0.03) |
| *3^rd^* | -0.29 (-1.14 to 0.57) | -0.44 (-1.42 to 0.55) |
| *4^th^* | -0.25 (-1.1 to 0.61) | -0.29 (-1.28 to 0.69) |
| *5^th^* | 0.48 (-0.36 to 1.32) | -0.1 (-1.07 to 0.86) |
| *6^th^* | 0 (-0.85 to 0.84) | -0.47 (-1.44 to 0.5) |
| *7^th^* | 0.21 (-0.63 to 1.04) | -0.36 (-1.32 to 0.6) |
| *8^th^* | 0.73 (-0.1 to 1.57) | 0.53 (-0.42 to 1.49) |
| *9^th^* | 0.88 (0.06 to 1.7) | 0.04 (-0.9 to 0.98) |
| *10^th^* | 0.57 (-0.24 to 1.38) | 0.16 (-0.77 to 1.09) |
|  | Overweight or obesity at age 10 years (%) | |
| *1^st^* | 7.45 (-9.28 to 24.18) | ref |
| *2^nd^* | 17.01 (0.47 to 33.54) | 10.7 (-8.04 to 29.44) |
| *3^rd^* | 19.71 (3.18 to 36.24) | 14.15 (-4.53 to 32.84) |
| *4^th^* | 19.06 (2.96 to 35.17) | 20.61 (2.15 to 39.12) |
| *5^th^* | 25.47 (9.38 to 41.57) | 21.63 (3.46 to 39.87) |
| *6^th^* | 31.36 (15.58 to 47.15) | 28.15 (10.06 to 46.32) |
| *7^th^* | 45.64 (29.99 to 61.3) | 32.06 (14.24 to 49.98) |
| *8^th^* | 41.26 (25.9 to 56.62) | 44.83 (27.26 to 62.53) |
| *9^th^* | 60.18 (45.21 to 75.14) | 48.39 (31.24 to 65.69) |
| *10^th^* | 7.45 (-9.28 to 24.18) | 57.82 (40.91 to 74.93) |

Multiple imputation analysis models were adjusted for maternal parity, maternal ethnicity, maternal and paternal BMI, maternal and paternal education, maternal and paternal smoking, maternal and paternal age, offspring sex, and cohort. Complete-case analysis models were adjusted for maternal parity, maternal ethnicity, maternal BMI, maternal education, maternal smoking, maternal age, offspring sex, and cohort.

### eFigure 1. Observed height, weight, and BMI in the Discovery and Replication cohorts

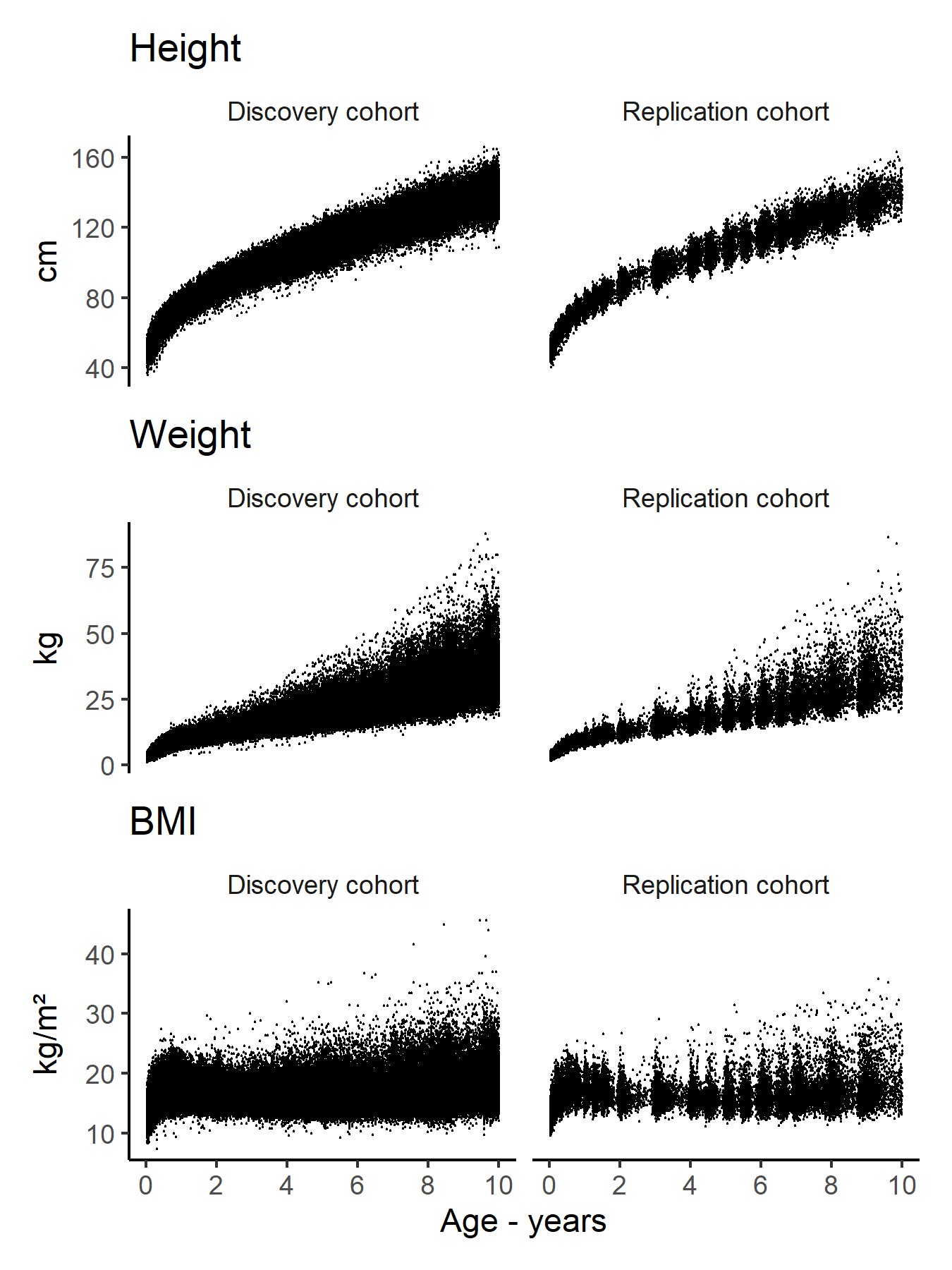

### eFigure 2. Predicted individual-specific growth trajectories in Discovery and Replication cohorts

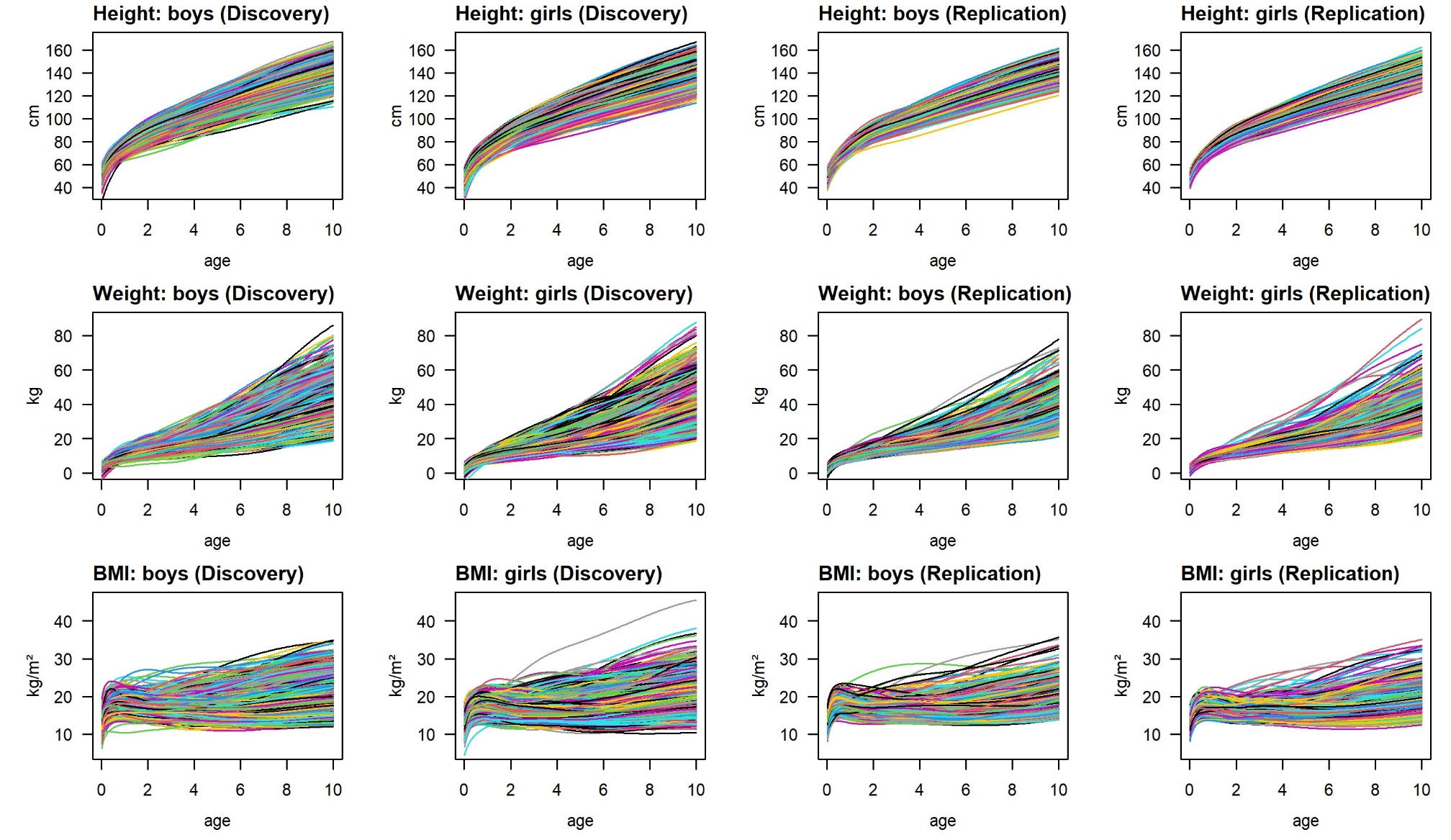

### eFigure 3. Correlations between infant/child growth outcomes in the European and replication birth cohorts

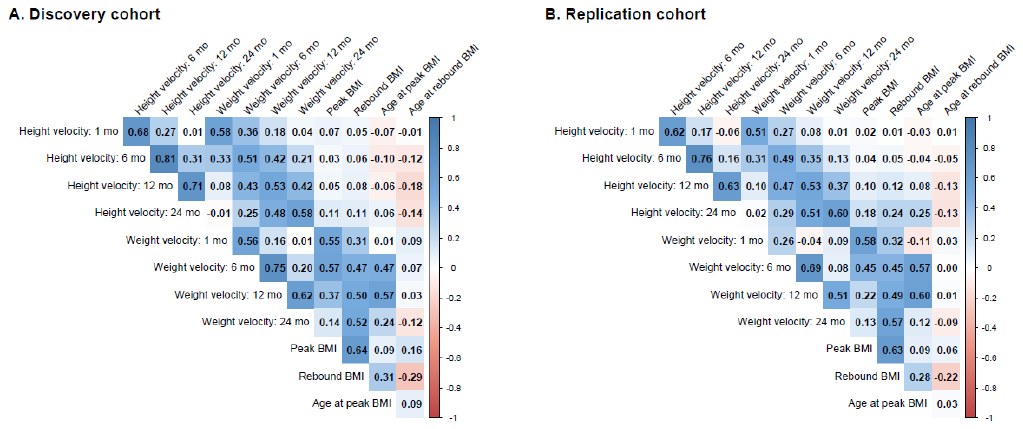

### eFigure 4. Mean height and weight Z score trajectory from age 1 month to 5 years, presented by sex in the Discovery cohort

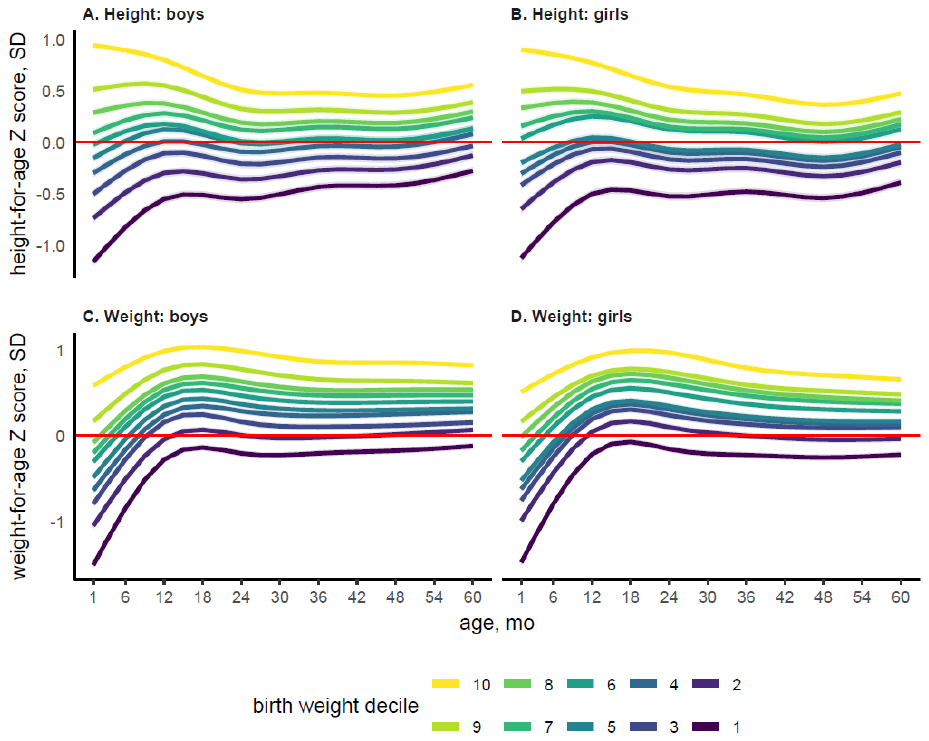

Estimates from models with a three-way interaction between age, birth weight, and sex and adjustment for cohort. Decile 1: <10^th^, decile 2: 10^th^ to <20^th^, decile 3: 20^th^ to <30^th^, decile 4: 30^th^ to <40^th^, decile 5: 40^th^ to <50^th^, decile 6: 50^th^ to <60^th^, decile 7: 60^th^ to <70^th^, decile 8: 70^th^ to <80^th^, decile 9: 80^th^ to 90^th^, decile 10: >90^th^

### eFigure 5. Nonlinear association of continuous birth weight Z-score with infant/child growth outcomes in the Discovery cohort

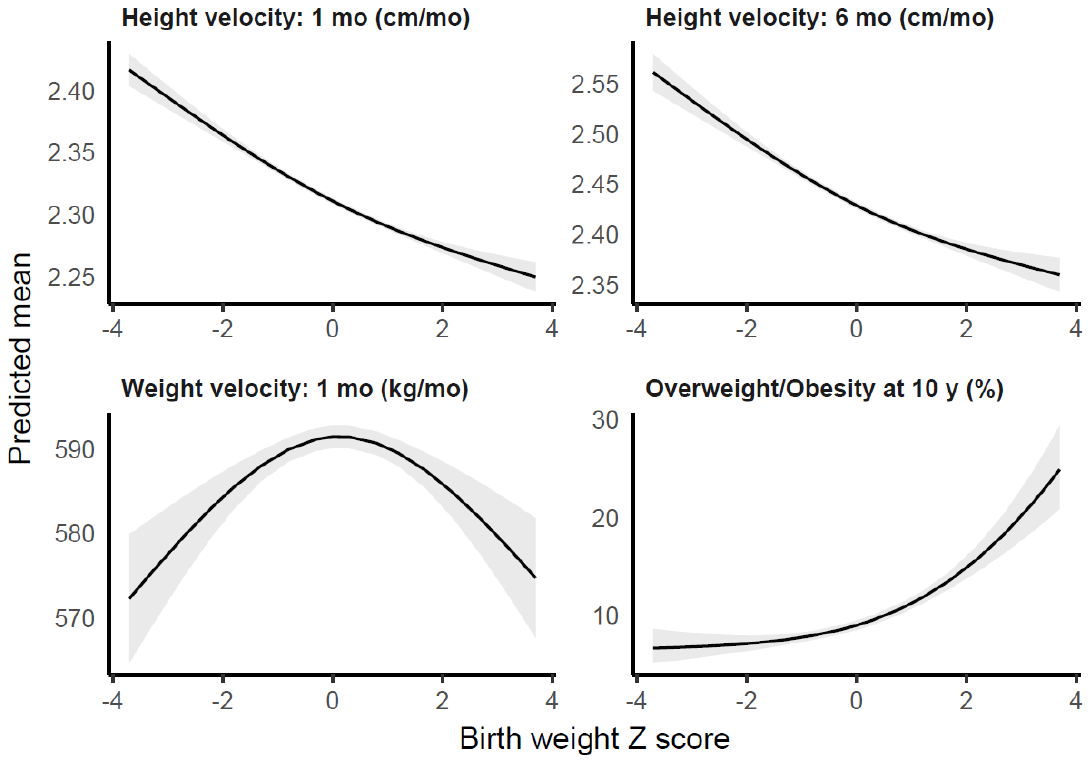

### eText 1. Cohort-specific acknowledgements and funding

**ABCD**

We are grateful to all participating hospitals, obstetric clinics, and general practitioners for their assistance in implementing the ABCD study and thank all of the women who participated for their cooperation. Core funding of the ABCD-study is provided by the Academic Medical Centre, Amsterdam, the Public Health Services, Amsterdam, and the Dutch Organization for Health Research and Development (ZonMw).

**ALSPAC**

We are extremely grateful to all of the families who took part in ALSPAC, the midwives for their help in recruiting them, and the whole ALSPAC team, which includes interviewers, computer and laboratory technicians, clerical workers, research scientists, volunteers, managers, receptionists and nurses. Core funding for the Avon Longitudinal Study of Parents and Children (ALSPAC) is provided by the UK Medical Research Council and Wellcome (217065/Z/19/Z) and the University of Bristol. A comprehensive list of grants funding is available on the ALSPAC website (http://www.bristol.ac.uk/alspac/external/documents/grant-acknowledgements.pdf). The funders had no role in the design of the study, the collection, analysis, or interpretation of the data; the writing of the manuscript, or the decision to submit the manuscript for publication. The views expressed in this paper are those of the authors and not necessarily those of any funder.

**BiB**

BiB receives core funding from the Wellcome Trust (WT101597MA and 223601/Z/21/Z), a joint grant from the UK Medical and Economic and Social Science Research Councils (MR/N024397/1), British Heart Foundation (CS/16/4/32482), and the National Institute of Health Research under its Applied Research Collaboration for Yorkshire and Humber (NIHR200166) and Clinical Research Network research delivery support. BiB is only possible because of the enthusiasm and commitment of the Children and Parents in BiB. We are grateful to all the participants, teachers, school staff, health professionals and researchers who have made BiB happen.

**EDEN**

The authors thank the cohort participants and the EDEN mother-child study group, whose members are: I. Annesi-Maesano, J.Y. Bernard, J. Botton, M.A. Charles, P. Dargent-Molina, B. de Lauzon-Guillain, P. Ducimetière, M. de Agostini, B. Foliguet, A. Forhan, X. Fritel, A. Germa, V. Goua, R. Hankard, B. Heude, M. Kaminski, B. Larroque†, N. Lelong, J. Lepeule, G. Magnin, L. Marchand, C. Nabet, F Pierre, R. Slama, M.J. Saurel-Cubizolles, M. Schweitzer, O. Thiebaugeorges. The EDEN study was supported by Foundation for medical research (FRM), National Agency for Research (ANR), National Institute for Research in Public health (IRESP: TGIR cohorte santé 2008 program), French Ministry of Health (DGS), French Ministry of Research, INSERM Bone and Joint Diseases National Research (PRO-A) and Human Nutrition National Research Programs, Paris-Sud University, Nestlé, French National Institute for Population Health Surveillance (InVS), French National Institute for Health Education (INPES), the European Union FP7 programmes (FP7/2007- 2013, HELIX, ESCAPE, ENRIECO, Medall projects), Diabetes National Research Program (through a collaboration with the French Association of Diabetic Patients (AFD)), French Agency for Environmental Health Safety (now ANSES), Mutuelle Générale de l’Education Nationale a complementary health insurance (MGEN), French national agency for food security, French speaking association for the study of diabetes and metabolism (ALFEDIAM).

**Generation XXI**

The Generation XXI was funded by Programa Operacional de Saúde – Saúde XXI, Quadro Comunitário de Apoio III and Administração Regional de Saúde Norte (Regional Department of Ministry of Health).  The Generation XXI is coordinated and conducted by several research groups from the Epidemiology Research Unit (EPIUnit) of the Institute of Public Health of the University of Porto (ISPUP), which was supported by FCT - Fundação para a Ciência e Tecnologia, I.P. through the projects with references UID/4750/2025 and LA/P/0064/2020 and DOI identifiers <https://doi.org/10.54499/UID/04750/2025> and <https://doi.org/10.54499/LA/P/0064/2020>. Susana Santos was supported by the European Union´s Horizon Europe Research and Innovation Programme under the Marie Sklodowska-Curie Postdoctoral Fellowship Grant Agreement No. 101109136 (URBANE). Views and opinions expressed are however those of the authors only and do not necessarily reflect those of the European Union or the European Research Executive Agency (REA). Neither the European Union nor the granting authority can be held responsible for them. We gratefully acknowledge the families enrolled in Generation XXI for their kindness, the participating hospitals and their staff for their help and support, and all previous and current members of the research and field team for their enthusiasm and perseverance.

**Project Viva**

We are indebted to the Project Viva mothers, children and families for their ongoing participation. The Project Viva study is supported by the US National Institutes of Health (R01HD034568). Project Viva and its team of co-investigators have been funded by grants from the National Institutes of Health (R01 HD 034568, R01 HL 075504, R01 HL 64925, R01 MH 068596, R01 HD 034568, R01 HD 064925, R01 ES 016314, U54 CA155626, R37 HD 034568, R01 AI 102960, R03 DE14004, P30 ES000002, R21 DK 073739, R21 DK 082661, R01 MD 003963, RC1 HD 063590, R01 HD 034568-0951, R01 ES21447, R01 NR013945, R01 HL111108, K24 HL 068041, R01 HL 64925, K23 HD 044807, T32 DK 081505, K23 DK083817, K24 HD069408, K12 DK094721), the Centers for Disease Control and Prevention (200- 95-0957), the Environmental Protection Agency (RD83479801), and the March of Dimes Foundation, with additional support from the Harvard Pilgrim Health Care Foundation.

**GUSTO**

We thank the GUSTO study group and all clinical and home-visit staff involved. The voluntary participation of all participants is greatly appreciated. The GUSTO study group includes. Allan Sheppard,Amutha Chinnadurai, Anne Ferguson-Smith, Anne Eng Neo Goh, Arijit Biswas, Audrey Chia, Birit Leutscher-Broekman, Borys Shuter, Shirong Cai, Cheryl Ngo, Chai Kiat Chng, Shang Chee Chong, Christiani Jeyakumar Henry, Mei Chien Chua, Cornelia Yin Ing Chee, Yam Thiam Daniel Goh, Dennis Bier, Chun Ming Ding, Doris Fok, Eric Andrew Finkelstein, Fabian Kok Peng Yap, George Seow Heong Yeo, Wee Meng Han, Helen Chen, Hugo P S Van Bever, Hazel Inskip, Iliana Magiati, Inez Bik Yun Wong, Jeevesh Kapur, Jenny L Richmond, Jerry Kok Yen Chan, Joshua J Gooley, Krishnamoorthy Niduvaje, Bee Wah Lee, Yung Seng Lee, Leher Singh, Sok Bee Lim, Lourdes Mary Daniel, Seong Feei Loh, Yen-Ling Low, Pei-Chi Lynette Shek, Marielle Fortier, Mark Hanson, Mary Foong-Fong Chong, Michael Meaney, Susan Morton, Wei Wei Pang, Pratibha Agarwal, Anqi Qiu, Boon Long Quah, Rob M van Dam, David Stringer, Salome Antonette Rebello, Wing Chee So, Chin-Ying Hsu, Lin Lin Su, Jenny Tang, Kok Hian Tan, Soek Hui Tan, Oon Hoe Teoh, Victor Samuel Rajadurai, PC Wong and Sudhakar K Venkates.
